# The Externalizing Spectrum and Suicide Risk: Insights from Genomics and Electronic Health Records in over 500,000 Veterans

**DOI:** 10.1101/2025.08.19.25332327

**Authors:** Peter B. Barr, Mallory Stephenson, Holly E. Poore, Chris Chatzinakos, Maissa Trabilsy, Xuejun Qin, Allison E. Ashley-Koch, Elizabeth R. Hauser, Jacquelyn L. Meyers, Roseann E. Peterson, Sandra Sanchez-Roige, Travis T. Mallard, Danielle M. Dick, K. Paige Harden, Mihaela Aslan, Philip D. Harvey, Jean C. Beckham, Tim B. Bigdeli, Nathan A. Kimbrel, COGA Collaborators, Cooperative Studies Program (CSP) #572, VA Million Veteran Program (MVP)

**Author notes:** Corresponding author: Peter B. Barr, Department of Psychiatry and Behavioral Sciences, SUNY Downstate Health Sciences University, 450 Clarkson Ave, MSC 1203, Brooklyn, NY 11203,. These authors jointly supervised this work (Tim B. Bigdeli, Nathan A. Kimbrel).

## Abstract

**Background:** Suicide-related outcomes, including suicidal ideation, suicide attempt, and suicide death, originate from heterogeneous mechanisms, including behavioral disinhibition characteristic of “externalizing” disorders (e.g., substance use disorders, ADHD, etc.). Prior work has demonstrated strong genetic overlap between externalizing and suicide attempts, suggesting this may provide valuable information about underlying suicide risk.

**Methods:** We investigated the co-occurrence between suicide-related outcomes and externalizing using data from the Million Veteran Program (MVP, N = 591,818) to: (1) create and validate a latent genomic factor for externalizing (MVP-EXT), (2) examine the genetic overlap between externalizing and suicidality using genetic correlations and polygenic scores (PGS), and (3) leverage the known genetic overlap to explore the prospective association between phenotypic externalizing and suicide death in electronic health records where measures of behavioral disinhibition are often lacking.

**Results:** The genetic correlation between MVP-EXT and a prior externalizing factor that did not contain MVP data was strong (rG = 0.87, 95% CI = 0.83, 0.91). MVP-EXT was genetically correlated with suicide attempt in veterans of European-like (rG = 0.67, 95% CI = 0.60, 0.74) and African-like (rG = 0.62, 95% CI = 0.42, 0.81) descent. PGSs were associated with suicidal ideation (OR = 1.09, 95% CI = 1.05, 1.13) and suicide attempts (OR = 1.20, 95% CI = 1.13, 1.27). An externalizing score derived from recent diagnoses (past 12-month at baseline) was prospectively associated with suicide death (HR = 1.36, 95% CI = 1.30, 1.42). The 5-year cumulative incidence for suicide death was ∼5x greater for those with 4+ past year externalizing codes compared to those with none.

**Conclusions:** Externalizing, reflective of impulsivity and behavioral disinhibition, is an important indicator of risk for suicide-related outcomes. The results here demonstrate externalizing and suicide may share a common genetic origin, in part, and this can be leveraged in electronic health records where measures of behavioral disinhibition are often non-existent. While the relation between suicide with internalizing disorders has generally received more attention, greater attention should be paid to externalizing as potential antecedents of suicide-related outcomes.

## Introduction

Suicide remains a leading preventable cause of death. Suicide rates in the United States increased ∼37% between 2000 and 2022. Deaths by suicide have contributed significantly to the decline in U.S. life expectancy, alongside other “deaths of despair” ^1,2^. The number of U.S. adults reporting a recent suicide attempt has increased from 0.62% in 2004-2005 to 0.79% in 2012-2013, with increases were even more pronounced among young adults and those with less education ^3^. Suicide attempts are more prevalent in those with co-occurring psychiatric problems. For example, among those with an alcohol use disorder (AUD), the prevalence of lifetime suicide attempt is three times that of the general population (17.5%) ^4^ and 40% of those seeking treatment for AUD report at least one suicide attempt ^5–8^. Additionally, meta-analyses of attention-deficit/hyperactivity disorders (ADHD) and suicide-related outcomes reveal strong associations with attempt, plans, and death by suicide ^9^. While mood-related problems are important risk factors for suicide present in these comorbid psychiatric problems ^10^, other mechanisms related to behavioral disinhibition and impulsivity are also important for understanding suicide risk. Unfortunately, widespread availability of measures for impulsivity and/or behavioral disinhibition are often limited in clinical settings or medical records. When patients have information on these domains, they have typically been measured in those being screened for other problems, biasing downstream associations ^11^ and limiting our ability to estimate any potential prognostic benefit.

Disorders related to behavioral disinhibition (e.g., substance use disorders, ADHD, etc.), generally referred to as externalizing disorders ^12,13^, share a common etiology ^14^, which is highly heritable ^15,16^, and overlaps strongly personality characteristics such as impulsivity ^17^. Recent multivariate genome wide association studies (GWAS) have found robust evidence for a shared genomic architecture for externalizing ^18–20^. This genetic liability to externalizing is genetically correlated with suicide attempt ^18,21^, and externalizing polygenic scores (PGS) are associated with suicide-related outcomes ^22,23^. Moreover, cases of suicide death were enriched for genomic risk for a variety of traits related to externalizing, including AUD and other mental health problems ^24^. However, we do not fully understand the nature of genetic overlap between externalizing and suicide-related outcomes.

In the current analysis, we leverage genomic and electronic health record (EHR) data to create and validate a measure of externalizing (as a proxy for behavioral disinhibition) in the Department of Veterans Affairs (VA) Million Veteran Program (MVP) cohort. Our approach starts with the shared molecular overlap of between externalizing and suicide-related outcomes, followed by an examination of the epidemiological relevance for externalizing and suicide-related mortality with the express purpose of improving identification of early risk indicators. Prior work has established that both cumulative risk scores of psychiatric (and externalizing) disorders ^25^ and substance use disorders ^26^ are predictive of suicidal behaviors (death and/or non-fatal attempt) in EHR. We expand on this work by 1) focusing on a narrower set of risk factors with a common genetic etiology and 2) suicide death, exclusively. First, we validated a latent genomic factor for externalizing in MVP (MVP-EXT), comparing results to a previously published genome wide association study (GWAS) ^18^. Next, compared the overlap of MVP-EXT and suicide-related outcomes using genetic correlations and polygenic scores in external cohorts. Lastly, we evaluated the prospective relationship between externalizing and suicide death from electronic health records (EHR) within MVP. In doing so, we create EHR-based measure of externalizing, validated on genetic data to explore the role externalizing may play in suicide death.

## METHODS

### The Million Veteran Program Cohort (MVP)

MVP links genomic laboratory testing, survey-based self-report data, and EHR data, with the goal of enhancing precision medicine initiatives ^27^. Enrolled participants reflect the population that utilizes the VA, with over-representation of older and male individuals, as well as higher rates of multiple morbidities and chronic conditions related to externalizing compared to the general population ^28,29^. Participants are active users of the VA healthcare system. Informed consent and authorization per the Health Insurance Portability and Accountability Act (HIPAA) were the only inclusion criteria. Once enrolled, participants’ EHR data are linked with their genetic data. The current analysis uses Release 4 of MVP data [February 2022] and was approved by the VA Central Institutional Review Board (IRB). All participants provided written informed consent.

MVP participants were genotyped on the MVP 1.0 Axiom array^30^. Genetic similarity of participants was classified using the HARE method ^31^, which harmonizes the closest inferred ancestral population with self-reported race and ethnicity. Genotypic data were imputed to the Trans-Omics for Precision Medicine (TOPMed) reference panel ^32^. We used data from the N = 467,101 veterans most similar to European reference panels (EUR-like) and the N = 124,717 veterans most similar to African reference panels (AFR-like) groups, as these had adequate statistical power for inclusion in the multivariate GWAS.

### Genome Wide Association Study (GWAS) Phenotypes

We identified externalizing-related traits within MVP data that matched those used in Externalizing Consortium GWAS (EXT1.0) ^18^ as closely as possible. Outcomes for the GWAS included in the multivariate models came from: 1) electronic health records (EHR) and 2) the MVP Baseline Survey. EHR data were converted to phecodes (version 1.2), which are clusters of ICD-9/10-CM codes ^33,34^. We defined a lifetime diagnosis for any given phecode as two or more occurrences of that phecode in their EHR, consistent with prior work ^35,36^. We examined all lifetime diagnoses without exclusion for overlap. We used phecodes for *Substance addiction and disorders* (Phecode 316, DUD), *Alcohol-related disorders* (Phecode 317, AUD), *Tobacco use disorder* (Phecode 318, TUD), and *Attention deficit hyperactivity disorder* (Phecode 313.1, ADHD). Lifetime smoking (SMOK) and binge drinking (BINGE) came from the Baseline Survey. We performed all univariate GWASs using SAIGE ^37^ to adjust for relatedness and included age, gender, and the first 20 genetic principal components as covariates (full details presented in the supplementary information). We filtered input GWAS to MAF > 1% with imputation scores (INFO) ≥ 0.80.

### Multivariate GWAS and downstream analyses

We performed a confirmatory factor analysis using GenomicSEM ^38^, which is robust to sample overlap and sample-size imbalances ^39^. Within the EUR-like population we used the provided linkage disequilibrium (LD) scores ^40^. For the AFR-like models, we used within-sample LD scores calculated using cov-LDSC ^41^. We carried the best fitting models forward for multivariate GWAS and ran the results from each population through FUMA (Functional Mapping and Annotation of GWAS) ^42^ to identify independent loci (*r*^2^ < 0.1) among genome-wide significant SNPs. Next, we estimated genetic correlations within GenomicSEM between the (latent) MVP-EXT factor, and observed indicators for suicide attempt ^24^, suicidal ideation ^43^. To further quantify the polygenic overlap between externalizing and suicide attempt, we applied bivariate MiXeR ^44^ which uses a bivariate Gaussian mixture model to estimate the proportion of shared influential genetic variance between two traits. Third, we used a series of genomic structural regression models to estimate the overlap of externalizing and suicide attempt in the presence of risk for other psychiatric disorders. Finally, we ran all results through a standard series of post-GWAS pipelines (detailed in the supplementary information).

### Independent validation cohorts

We included two independent cohorts to perform follow-up analyses. First, the Collaborative Study on the Genetics of Alcoholism (COGA) is a multi-site study of families densely affected with AUD and community-ascertained comparison families ^45–47^. Participants completed a poly-diagnostic interview, the Semi-Structured Assessment for the Genetics of Alcoholism (SSAGA) ^48^. The final analytic sample consisted of 10,986 COGA participants with genetic data ^49^ (N_EUR-like_ = 7,601; N_AFR-like_ = 3,385). Second, the National Longitudinal Study of Adolescent to Adult Health (Add Health) is a nationally representative study of participants in the United States recruited as adolescents and followed into adulthood ^50^. Our final analytic sample for Add Health consisted of 6,883 individuals (N_EUR-like_ = 5,122; N_AFR-like_ = 1,761). Both samples included information on suicide attempt and suicidal ideation. Individuals were coded as having lifetime suicidal ideation or a lifetime suicide attempt (not mutually exclusive) if they answered “Yes” to these questions at any point during data collection. Full descriptions of each cohort are presented in the supplemental information.

### Polygenic scores (PGS)

We estimated polygenic scores (PGS, aggregate measures of risk alleles weighted by GWAS effect sizes) using PRS-CSx ^51^, which integrates GWAS summary statistics across multiple populations to improve the predictive power of PGS in the populations that typically lack well-powered GWAS results. Because of variation in allele frequencies and LD, PGS lose predictive accuracy when there is mismatch between the population of the discovery GWAS and target sample, even within relatively homogenous clusters ^52^. PRS-CSx employs a Bayesian approach to correct GWAS summary statistics for the non-independence of SNPs in the genome. We converted PGS into Z-scores and included age, sex, the first 10 genetic principal components, and cohort specific covariates in all analyses.

### Prospective investigation of externalizing and suicide-related mortality in EHR

Lastly, to better contextualize externalizing in relation to prospective risk for death by suicide, we explored associations between externalizing and post-enrollment mortality. For each participant, we created a count of the past 12-month externalizing EHR codes (relative to MVP enrollment) and examined whether this count was associated with mortality, adjusting for competing risk of all other mortality models via Fine-Gray models ^53,54^. We used cause of death (COD) from the ICD codes provided in the linked National Death Index (NDI) data up to December 2021 (v21, see Supplemental Table 1 for specific codes). The competing risk model allowed us to accurately estimate the hazards for specific causes of death (e.g., suicide death) in the context of competing causes (e.g., all other causes of death). We explored two competing risk models: 1) suicide-death in the context of risk for all other causes of death, and 2) “deaths of despair” (suicide, alcohol, and drug related deaths) in the context of risk for all other causes of death. We included age, sex, and race-ethnicity (American Indian/Alaskan Native, Asian, Black/African American, Hispanic or Latino/a/x, multiracial, Native Hawaiian/Pacific Islander, non-Hispanic White, or other race or ethnicity) as covariates. Analyses were conducted using the *survival* package in *R* ^55,56^.

It is important to note this paper includes language related to both race-ethnicity, which reflects socially-constructed categories, and genetic similarity, which uses empirical assignment based on available reference panels, because both are relevant for the current analyses. Prior work has established that racism, discrimination, and adverse social conditions — to which marginalized populations are disproportionally exposed — are relevant to suicide outcomes ^57–59^. Additionally, we follow best practices for handling genetic data from diverse populations to limit bias from population stratification ^60^. The inclusion of both concepts in no way endorses the notion that these reflect discrete biological categories.

## Results

### Creating and validating a latent externalizing factor in MVP

Table 1 presents the individual GWAS results for the 6 indicators in the multivariate GWAS. Our final model for EUR-like veterans (N_eff_ = 310,498) contained all six indicators and showed reasonably good fit to this model specification (Figure 1, Panel A; *χ*^2^= 104.10, df = 7, p = 1.53×10^−19^; CFI = 0.99, SRMR = 0.06). For the AFR-like veterans, the genetic correlation between SMOK and TUD was indistinguishable from one. We therefore retained only TUD, as it had greater statistical power. We also replaced ADHD with the attention problems subscale of the Medical Outcomes Survey as a proxy (MOS-ATTN, *h*^2^_SNP_ [SE] = 0.033 [0.009], LDSC intercept = 1.003, Mean χ^2^ = 1.038, λ_GC_ = 1.035, rG with ADHD = 0.8) due to the low power of the ADHD indicator. The final model for AFR-like veterans (N_eff_ = 99,949) therefore, largely mirrored that of the EUR-like veterans with one fewer indicator (Figure 1, Panel B; *χ*^2^= 131.51, df = 8, p = 1.38×10^−24^; CFI = 0.96, SRMR = 0.09).

**Figure 1:**
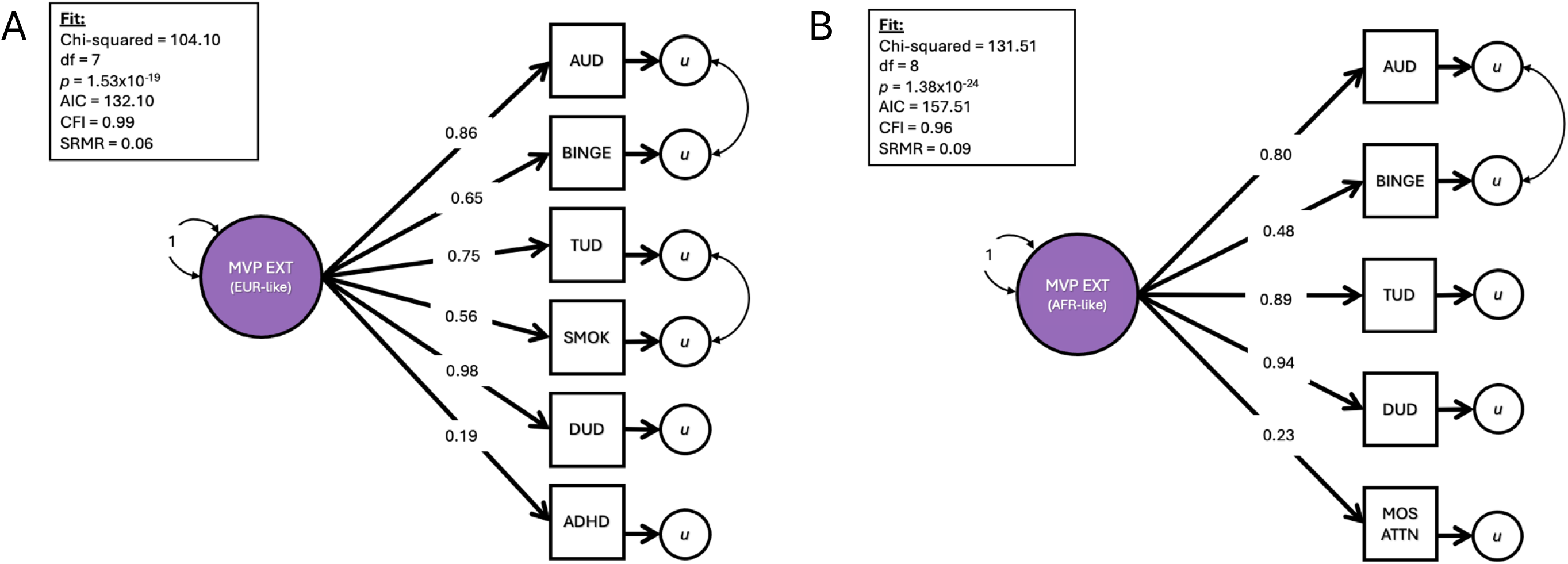
Multivariate GWAS models of externalizing in MVP. Final models for EUR-like (Panel A) and AFR-like (Panel B) populations used in the subsequent multivariate GWASs. *Df* = degrees of freedom. AIC = Akaike’s information criterion, CFI = comparative fit index, SRMR = standardized root mean squared residual.

**Table 1:**
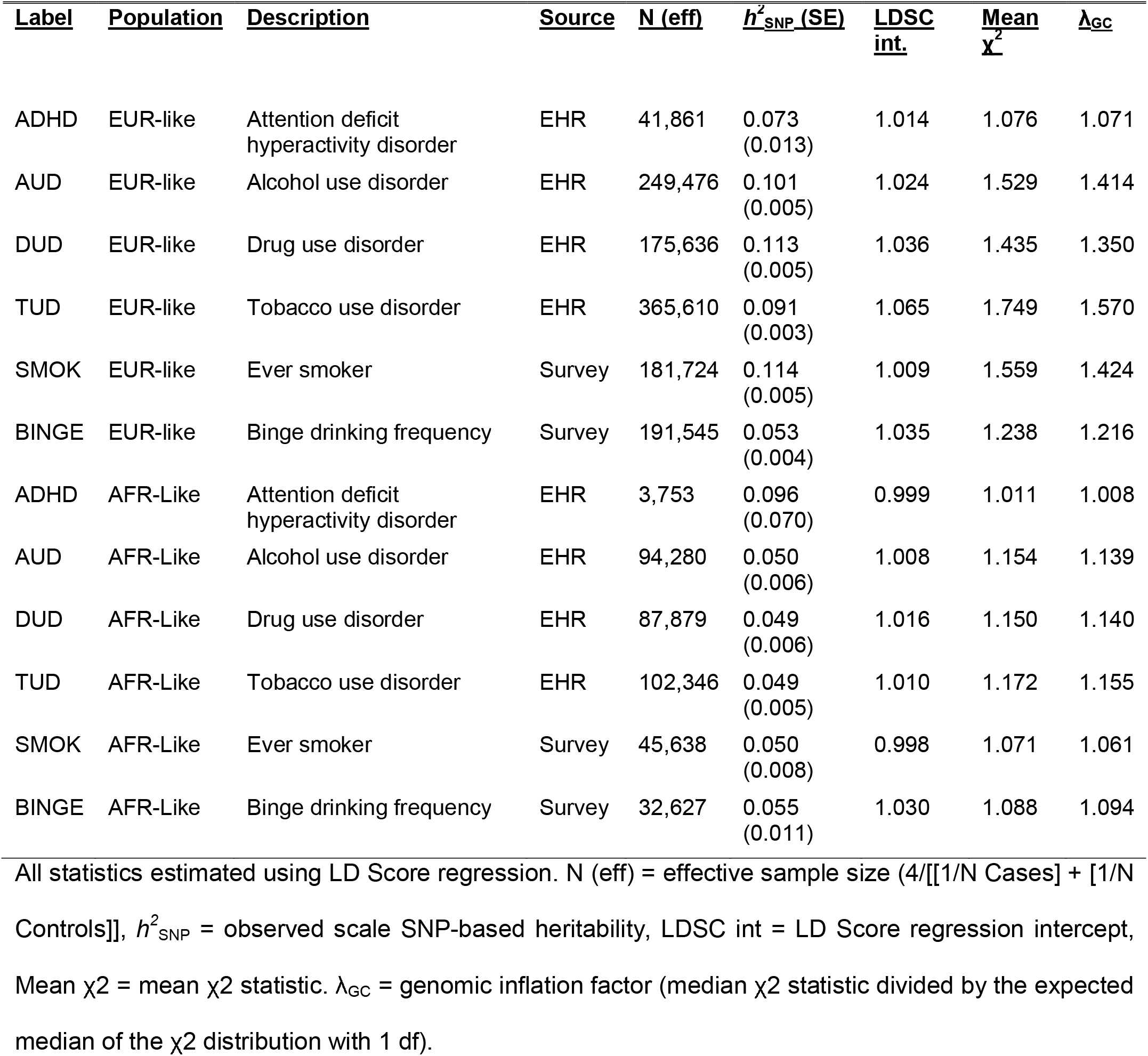
EXTERNALIZING PHENOTYPES INCLUDED IN THE MULTIVARIATE GWAS.

| <u>Label</u> | <u>Population</u> | <u>Description</u> | <u>Source</u> | <u>N (eff)</u> | <u>$h^2_{\text{SNP}}$ (SE)</u> | <u>LDSC int.</u> | <u>Mean $\chi^2$</u> | <u>$\lambda_{\text{GC}}$</u> |
| --- | --- | --- | --- | --- | --- | --- | --- | --- |
| ADHD | EUR-like | Attention deficit hyperactivity disorder | EHR | 41,861 | 0.073 (0.013) | 1.014 | 1.076 | 1.071 |
| AUD | EUR-like | Alcohol use disorder | EHR | 249,476 | 0.101 (0.005) | 1.024 | 1.529 | 1.414 |
| DUD | EUR-like | Drug use disorder | EHR | 175,636 | 0.113 (0.005) | 1.036 | 1.435 | 1.350 |
| TUD | EUR-like | Tobacco use disorder | EHR | 365,610 | 0.091 (0.003) | 1.065 | 1.749 | 1.570 |
| SMOK | EUR-like | Ever smoker | Survey | 181,724 | 0.114 (0.005) | 1.009 | 1.559 | 1.424 |
| BINGE | EUR-like | Binge drinking frequency | Survey | 191,545 | 0.053 (0.004) | 1.035 | 1.238 | 1.216 |
| ADHD | AFR-Like | Attention deficit hyperactivity disorder | EHR | 3,753 | 0.096 (0.070) | 0.999 | 1.011 | 1.008 |
| AUD | AFR-Like | Alcohol use disorder | EHR | 94,280 | 0.050 (0.006) | 1.008 | 1.154 | 1.139 |
| DUD | AFR-Like | Drug use disorder | EHR | 87,879 | 0.049 (0.006) | 1.016 | 1.150 | 1.140 |
| TUD | AFR-Like | Tobacco use disorder | EHR | 102,346 | 0.049 (0.005) | 1.010 | 1.172 | 1.155 |
| SMOK | AFR-Like | Ever smoker | Survey | 45,638 | 0.050 (0.008) | 0.998 | 1.071 | 1.061 |
| BINGE | AFR-Like | Binge drinking frequency | Survey | 32,627 | 0.055 (0.011) | 1.030 | 1.088 | 1.094 |

All statistics estimated using LD Score regression. N (eff) = effective sample size ( $4/[1/N \text{ Cases}] + [1/N \text{ Controls}]$ ), $h^2_{\text{SNP}}$ = observed scale SNP-based heritability, LDSC int = LD Score regression intercept, Mean $\chi^2$ = mean $\chi^2$ statistic. $\lambda_{\text{GC}}$ = genomic inflation factor (median $\chi^2$ statistic divided by the expected median of the $\chi^2$ distribution with 1 df).

Next, we performed multivariate GWASs within the EUR-like and AFR-like veterans, separately, followed by an inverse-variance weighted fixed effects meta-analysis in METAL ^61^. There were 155 independent genome wide significant loci in the meta-analysis, of which only 11 were significant Q-SNPs (*p* <.05/155 = 3.23 ×10^−4^). We took several steps to validate MVP-EXT with external data that did not contain MVP and included multiple non-SUD indicators. First, of the 138 loci present in EXT1.0 results, 94.2% (95% CI = 88.9%, 97.5%) were sign concordant and 116 (84.1%) of the loci were nominally significant (p < .05). Second, the genetic correlation between MVP-EXT and EXT1.0 was strong (rG = 0.87, 95% CI = 0.83, 0.91). Third, the genetic correlations of MVP-EXT and EXT1.0 with 92 external traits was strong (*r* = 0.96, 95% CI = 0.93, 0.97). Lastly, the MVP-EXT PGS was associated with a matched latent factor of externalizing in COGA and Add Health, as well as on overall meta-analyzed effect (Beta *_META_* = 0.20, 95% CI = 0.18, 0.22). Taken together, these results suggest that the MVP-EXT factor largely recapitulated EXT1.0 (full results from GWAS and post-GWAS biological characterization presented in the supplementary information and Supplemental Tables 2-8).

### Exploring the genomic overlap between externalizing and suicide related outcomes

We next characterized the genetic overlap between MVP-EXT and suicide related phenotypes. First, we estimated the genetic correlations between MVP-EXT and both suicide attempt (SA) ^62^ and suicidal ideation (SI) ^43^. Figure 2 (Panel A) presents genetic correlation estimates for the EUR-like and AFR-like results side by side. Genetic correlations with SA were strong for both the EUR-like (rG = 0.67, 95% CI = 0.60, 0.91) and AFR-like (rG = 0.74, 95% CI = 0.42, 0.81) participants. Genetic correlations with SI were weak, but significant in the EUR-like (rG = 0.12, 95% CI = 0.08, 0.17) veterans and null for AFR-like (rG = −0.01, 95% CI = −0.20, 0.18) veterans. The results from MiXeR indicated that MVP-EXT shared ∼77% of its influential variants with suicide attempt. Within this shared component, the variants that influence both MVP-EXT and suicide attempt had high sign concordance between SNPs (79%, SE = 0.03), supporting a model in which externalizing and suicide attempt were distinct traits with substantial polygenic overlap (full results in Supplemental Table 9). Finally, we compared the change in effect size for MVP-EXT and SA before and after covarying for depression ^63^ and schizophrenia ^64^ via genomic structural regression (Figure 2C). The association between MVP-EXT and SA is reduced by ∼50% when covarying for depression and schizophrenia, but remains significant (Beta = 0.285, SE = 0.04, *p =* 4.60×10^−12^).

**Figure 2:**
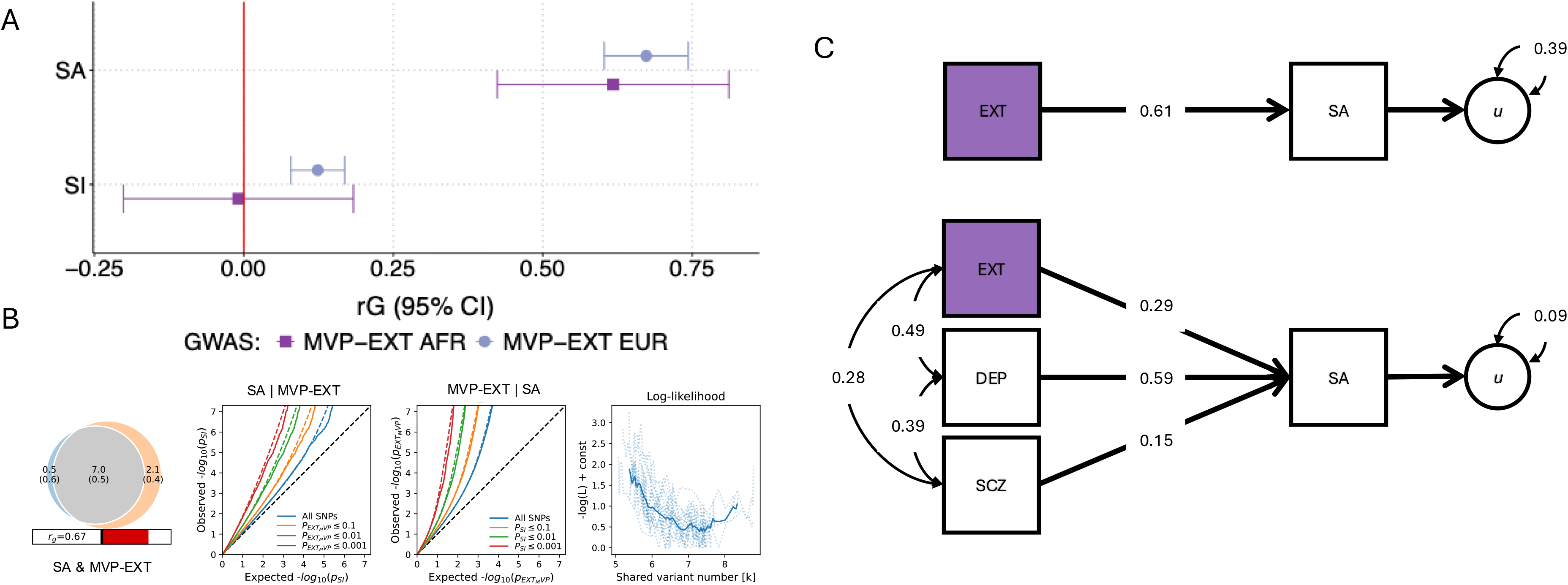
Genetic correlations across populations. Genetic correlations (and 95% confidence intervals) between MVP-EXT and suicide attempt (SUI) and suicidal ideation (IDE) for both European-like and African-like results (Panel A). Genetic overlap between MVP-EXT and SUI using bivariate MiXeR in European-like results (Panel B). Venn diagram depicting the estimated number of influencing variants in thousands shared (grey) between and unique to suicide attempt (SUI, blue) and externalizing (MVP-EXT, orange). Conditional quantile–quantile (Q–Q) plots show observed versus expected −log10 p values in SUI and MVP-EXT as a function of the significance of association with the other trait. Log likelihood curves show goodness of model fit as a function of the count of shared influential variants. Genomic structural regression models (Panel C) comparing models association between MVP-EXT and SA before and after covarying for depression and schizophrenia. All estimates are standardized.

Next, we examined the association between MVP-EXT PGS and suicide outcomes (full results in Supplemental Table 10). The MVP-EXT PGS was positively associated with lifetime SI and SA in EUR-like participants from both COGA (SI: OR = 1.18, 95% CI = 1.11, 1.25; SA: OR = 1.46, 95% CI = 1.32, 1.61) and Add Health (SI: OR = 1.07, 95% CI = 1.01, 1.13; SA: OR = 1.13, 95% CI = 1.03, 1.24), but not the AFR-like participants in either COGA (SI: OR = 0.98, 95% CI = 0.89, 1.08; SA: OR = 1.07, 95% CI = 0.94, 1.22) or Add Health (SI: OR = 1.04, 95% CI = 0.93, 1.16; SA: OR = 0.97, 95% CI = 0.82, 1.15). For both SI and SA, the meta-analyzed estimate was significant (SI: OR = 1.09, 95% CI = 1.05, 1.13; SA: OR = 1.20, 95% CI = 1.13, 1.27). For context, Figure 3, Panel B presents the proportion of respondents in COGA reporting a lifetime suicide attempt across deciles of 1) the MVP-EXT PGS and 2) EXT-factor scores derived from the phenotypes used for replication. We see a similar pattern across predictor (PGS vs factor scores) and population, whereby the associations are stronger in EUR-like participants.

**Figure 3:**
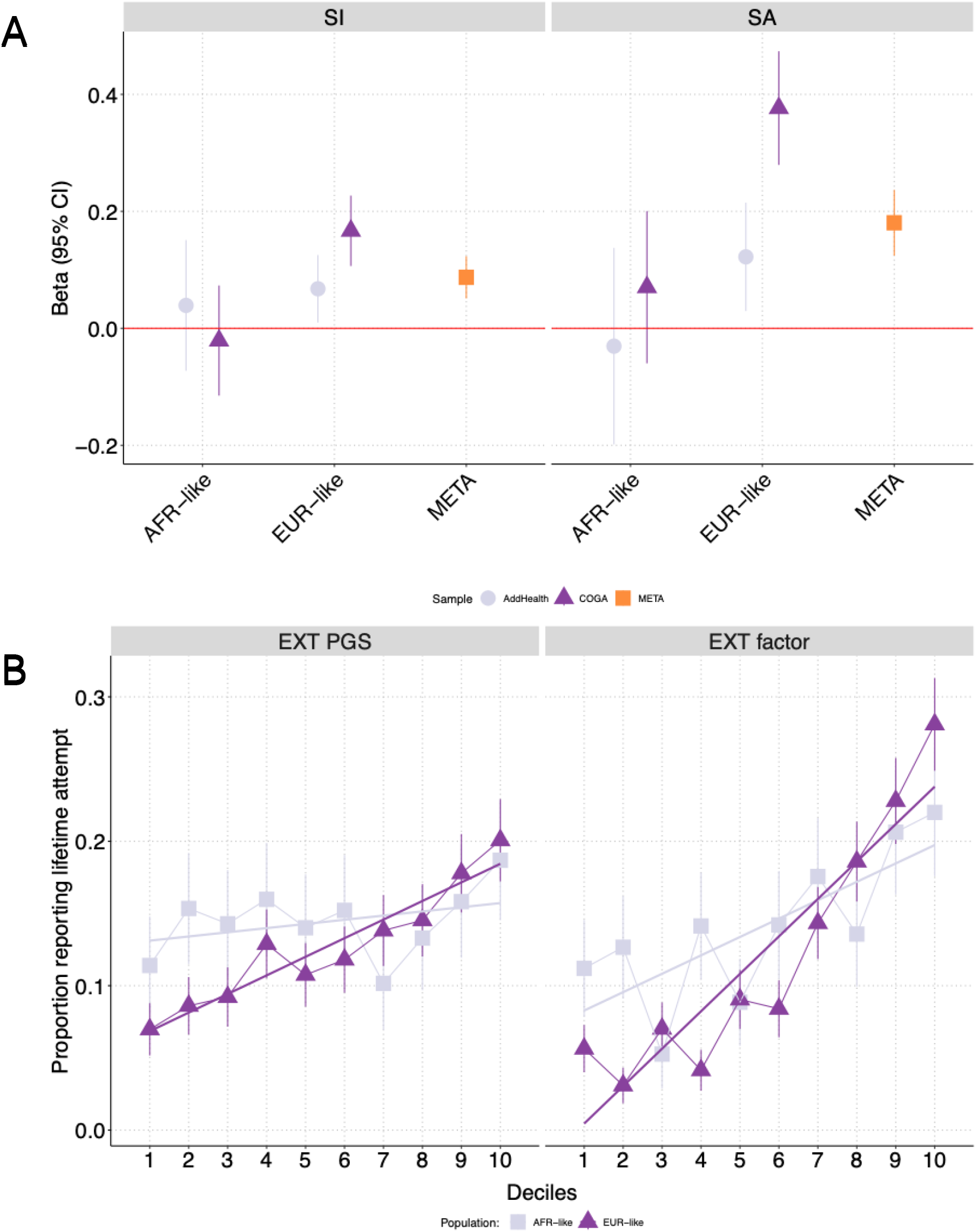
Polygenic associations between MVP-EXT PGS in COGA and Add Health. European-like, African-like, and meta-analysis associations between MVP-EXT PGS and suicide attempt (SA) and suicidal ideation (SI) in COGA and Add Health (Panel A). Proportion of individuals reporting lifetime suicide attempt across MVP-EXT PGS deciles (left) and phenotypic factor score deciles (right) in European-like and African-like COGA participants (Panel B).

### Prospective phenotypic analyses in electronic health records

Finally, we created counts of past 12-month externalizing codes from the time of MVP enrollment (N = 840,865 as of 12/31/2021, overlapping with NDI time frames). Demographic information is presented in Supplemental Table 11. Of these participants, 13.8% (N = 116,365) were deceased by the end of 2021. Of the deceased, N = 1,268 were classified as suicides, while N = 4,410 of the deaths met the more expansive definition for deaths of despair (which includes suicide). The results from the competing risk models are presented in Table 2. Externalizing codes were associated with both suicide death (HR = 1.360, 95% CI = 1.302, 1.420) and the broader deaths of despair category (HR = 1.936, 95% CI = 1.903, 1.970), even in the case of competing risk for all other causes of mortality. Additionally, there is a non-linear increase in the 5-year cumulative incidence for both suicide deaths and deaths of despair across count of EXT codes. For those with 4+ past year externalizing codes (compared to those with none), the 5-year cumulative incidence for suicide death was nearly 5x greater and the 5-year cumulative incidence for deaths of despair was almost 20x greater. As a final robustness check, we also covaried for past year diagnoses of major depression (phecode 296.22) and PTSD (phecode 300.9). The results were virtually unchanged suggesting that the association between EXT burden and suicide death was not explained by the increased rates of comorbid psychiatric conditions in those with EXT-related diagnoses.

**Table 2:**
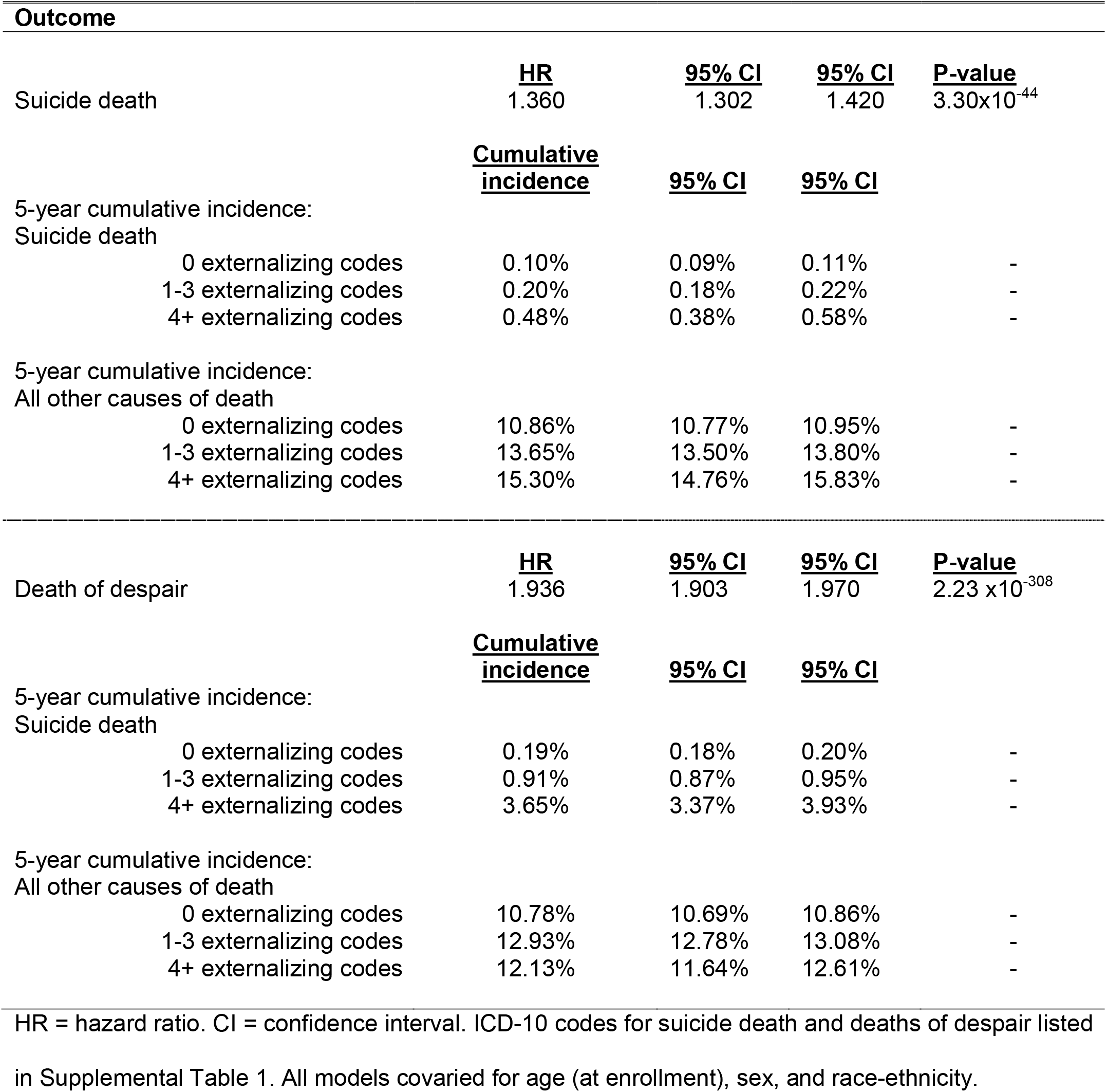
COMPETING RISK MODELS AND 5-YEAR CUMULATIVE INCIDENCE FOR COUNT OF EXTERNALIZING DIAGNOSES AND MORTALITY.

## Discussion

Externalizing has been consistently linked to suicide-related outcomes at the phenotypic ^22,65–67^ and genetic level ^18,22,23,68,69^, pointing to the relevance of behavioral disinhibition. In the current analysis, we sought to further explore this relationship in order to create a validated measure of externalizing for prospective analyses with suicide death. First, recapitulated a latent genetic externalizing factor that overlapped strongly with prior results (*rg* = .87) containing no MVP data ^18^. Second, we examined the shared molecular basis of genetic liability for externalizing and suicide-related outcomes using genetic data from European-like and African-like populations in the Million Veteran Program Cohort, again finding a similar pattern of results across the MVP and non-MVP derived externalizing factors. Finally, we explored the prognostic relevance of externalizing and suicide death by examining the prospective association between recent externalizing diagnoses and mortality in the full MVP sample.

In line with previous research, we found consistent evidence for genetic overlap between externalizing and suicide attempt. Genetic correlations between MVP-EXT and suicide-related outcomes across the EUR-like and AFR-like results had similar patterns of associations. MVP-EXT was strongly associated with suicide attempt in both populations, and results from MiXeR demonstrated that these are distinct phenotypes that have significant polygenic overlap. Importantly, the results from the genomic structural regression models demonstrated that, while attenuated, the association between externalizing and SA remained significant when covarying for depression and schizophrenia. This mirrors results from polygenic score analyses in MVP which found that the association between the externalizing PGS and the phecode for suicidal thought and behaviors remained significant even when covarying for depression, schizophrenia, and suicide attempt PGSs ^23^.

For suicidal ideation, MVP-EXT was significantly associated with suicidal ideation in the EUR-like results, though the confidence intervals overlapped with the AFR-like results, suggesting that the null association may be the result of the lower power. In the polygenic score analyses, the association between MVP-EXT PGS and suicide-related outcomes was null for the AFR-like participants. There are several possible explanations for this lack of association. First, it is possible we did not have sufficient power, as seen in the genetic correlation results. Second, our results are primarily derived from EHR. Marginalized racial and ethnic populations receive less engagement in EHR (e.g., fewer number of EHR actions performed within a patient’s EHR per hour) compared to those who are non-Hispanic White ^70^, leading to a potentially biased assessment of externalizing. Lastly, in line with the “social push” model ^71^ of gene-environment interaction, the null association could reflect the increased relevance of adverse social conditions amongst AFR-like participants (e.g., racism, discrimination) ^57–59^, who are predominantly African-American. Importantly, these are not mutually exclusive explanations.

Finally, beyond genetic analyses, which may capture more trait-like (e.g., aggerate lifetime risk) associations, current – as of time of enrollment – externalizing codes were associated with suicide related mortality, potentially capturing time-dependent risk. Prior work has found that cumulative burden of psychiatric, substance use, and other medical disorders over time are predictive of suicidal behaviors, including both attempt and death ^25^. Our analyses add to this literature by demonstrating that current externalizing diagnoses, specifically, are relevant for suicide death in particular. Importantly, the association with MVP-EXT and suicide death remains in the context of risk for other causes of death and comorbid diagnoses. Given the multitude of negative health correlates of EXT found in previous analyses of MVP (e.g., cirrhosis, chronic airway obstruction, viral hepatitis C, etc.)^23^ these results help frame the clinical importance of externalizing beyond the specific health consequences related to substance use. Development and implementation of externalizing risk screeners at intake may help to mitigate a variety of negative health outcomes, including suicide.

This research has several important limitations. First, while we utilized the largest sample of non-European participants available in MVP, we still lacked sufficient power within the AFR-like participants to identify loci associated with externalizing. As future releases of MVP become available, it will be important to include larger samples and data from additional populations. Second, these analyses focused solely on the role of genetic risk. Suicide-related outcomes are complex, and social factors (e.g., unemployment, experiences of homelessness, social support) also play an important role ^57^. Future efforts should examine genomic risk factors in conjunction with well-established social and clinical determinants of suicide-related outcomes, as has been done with SUD ^72^. Third, our sample was predominantly comprised of men (∼85%) and there are large gender differences in suicide-related outcomes, especially in regard to suicide attempt vs death. Whether the results from the current analysis generalize broadly remains an open question.

Suicide remains a critical public health problem with a complex etiology. While the relation between suicide and mood-related disorders has received greater attention, externalizing problems are an important factor in risk for suicide related behaviors. We recapitulated findings from a previous GWAS of externalizing problems using data derived exclusively from MVP. Our analysis demonstrates that externalizing risk is broadly associated with suicide-related outcomes, though it seems more relevant for suicide attempt and suicide death than suicidal ideation. Importantly, current diagnoses of externalizing problems are associated with future suicide death. Efforts at early intervention and suicide screening should consider addressing problems related to externalizing and behavioral disinhibition in addition to other common suicide-related risk factors.

## Acknowledgements

This research is based on data from the Million Veteran Program, Office of Research and Development, Veterans Health Administration, and was supported by MVP000. This work was also funded by grant #1I01CX001729 from the Clinical Services Research & Development (CSRD) Service of VA ORD to Drs. Beckham and Kimbrel, by the MVP CHAMPION program, which is a collaboration between the VA and the Department of Energy (DoE), and by a CSRD Senior Research Scientist award (#lK6BX003777) to Dr. Beckham. Dr. Kimbrel was also supported by a VA Research Career Scientist Award (#I01BX005881) from VA ORD. Drs. Barr, Bigdeli, Aslan, and Harvey are supported by VA Cooperative Studies Program (CSP) #572. Drs. Peterson, Bigdeli, and Meyers are supported by the National Institute of Mental Health (R01MH125938). Dr. Peterson is also supported by the National Institute on Alcohol Abuse and Alcoholism (P50AA022537) and the Brain Behavior Research Foundation NARSAD grant 28632 PS Fund. Drs. Barr and Dick are also supported by the National Institute of Drug Abuse (R01DA050721) and the National Institute of Alcohol Abuse and Alcoholism (R01AA015416). Dr. Sanchez-Roige was supported by funds from the California Tobacco-Related Disease Research Program (TRDRP; Grant Number T32IR5226) and the National Institute on Drug Abuse (DP1DA054394). Dr. Mallard is supported by funds from NIH T32HG010464. This publication does not represent the views of the Department of Veteran Affairs (VA), the National Institutes of Health, or the United States Government. Dr. Barr had full access to all the data in the study and takes responsibility for the integrity of the data and the accuracy of the data analysis. The funders had no role in the design and conduct of the study; collection, management, analysis, and interpretation of the data; preparation of the manuscript; and decision to submit the manuscript for publication. The MVP Publication Committee reviewed and approved the manuscript. A preprint of this manuscript was posted to medRxiv (doi: 10.1101/2025.08.19.25332327).

The Collaborative Study on the Genetics of Alcoholism (COGA), Principal Investigators B. Porjesz, V. Hesselbrock, A. Agrawal; Scientific Director, A. Agrawal; Translational Director, D. Dick, includes ten different centers: University of Connecticut (V. Hesselbrock); Indiana University (H.J. Edenberg, T. Foroud, Y. Liu, M.H. Plawecki); University of Iowa Carver College of Medicine (S. Kuperman, A. Anderson); SUNY Downstate Health Sciences University (B. Porjesz, J. Meyers); Washington University in St. Louis (L. Bierut, A. Agrawal, S. Hartz); University of California at San Diego (M. Schuckit); Rutgers University (D. Dick, R. Hart, J. Salvatore, J. Tischfield); The Children’s Hospital of Philadelphia, University of Pennsylvania (L. Almasy); Icahn School of Medicine at Mount Sinai (A. Goate, P. Slesinger); and Howard University (D. Scott). Other COGA collaborators include: C. Holzhauer, M. Hesselbrock (University of Connecticut); D. Lai, J. Nurnberger Jr., L. Wetherill, X., Xuei, S. O’Connor, (Indiana University); J. Kramer (University of Iowa), G. Chan (University of Iowa; University of Connecticut); C. Kamarajan, A. Pandey, D.B. Chorlian, P. Barr, S. Kinreich, G. Pandey, Z. Neale, S., C. Chatzinakos, J. Zhang, Saenz deViteri, R. Christian, A. Bingly (SUNY Downstate); G. Pathak (Icahn School of Medicine at Mount Sinai); A. Anokhin, K. Bucholz, F. Dong, A. Hatoum, E. Johnson, V. McCutcheon, J. Rice, S. Saccone (Washington University); F. Aliev, Z. Pang, S. Kuo, S. Brislin, J. Moore (Rutgers University); A. Merikangas (The Children’s Hospital of Philadelphia and University of Pennsylvania); M. Gitik, NIAAA Staff Collaborator. We continue to be inspired by our memories of Henri Begleiter and Theodore Reich, founding PI and Co-PI of COGA, and also owe a debt of gratitude to other past organizers of COGA, including Ting-Kai Li, P. Michael Conneally, Raymond Crowe, and Wendy Reich, for their critical contributions. This national collaborative study is supported by NIH Grant U10AA008401 from the National Institute on Alcohol Abuse and Alcoholism (NIAAA) and the National Institute on Drug Abuse (NIDA).

We would also like to thank The Externalizing Consortium for sharing the GWAS summary statistics of externalizing. The Externalizing Consortium: Principal Investigators: Danielle M. Dick, Philipp Koellinger, K. Paige Harden, Abraham A. Palmer. Lead Analysts: Richard Karlsson Linnér, Travis T. Mallard, Peter B. Barr, Sandra Sanchez-Roige. Significant Contributors: Irwin D. Waldman. The Externalizing Consortium has been supported by the National Institute on Alcohol Abuse and Alcoholism (R01AA015416 - administrative supplement), and the National Institute on Drug Abuse (R01DA050721). Additional funding for investigator effort has been provided by K02AA018755, U10AA008401, P50AA022537, as well as a European Research Council Consolidator Grant (647648 EdGe to Koellinger). The content is solely the responsibility of the authors and does not necessarily represent the official views of the above funding bodies. The Externalizing Consortium would like to thank the following groups for making the research possible: 23andMe Inc., Add Health, Vanderbilt University Medical Center’s BioVU, Collaborative Study on the Genetics of Alcoholism (COGA), the Psychiatric Genomics Consortium’s Substance Use Disorders working group, UK10K Consortium, UK Biobank, and Philadelphia Neurodevelopmental Cohort. We would like to thank the many studies that made these consortia possible, the researchers involved, and the participants in those studies, without whom this effort would not be possible. We would also like to thank the research participants and employees of 23andMe.

This research uses data from Add Health, funded by grant P01 HD31921 (Harris) from the Eunice Kennedy Shriver National Institute of Child Health and Human Development (NICHD), with cooperative funding from 23 other federal agencies and foundations. Add Health is currently directed by Robert A. Hummer and funded by the National Institute on Aging cooperative agreements U01 AG071448 (Hummer) and U01AG071450 (Aiello and Hummer) at the University of North Carolina at Chapel Hill. Add Health was designed by J. Richard Udry, Peter S. Bearman, and Kathleen Mullan Harris at the University of North Carolina at Chapel Hill.

Most importantly, we would like to thank the U.S. Veteran participants in MVP for their service, and for their time, samples, and continued participation in VA research. Without them, this work would not be possible.

## Conflicts of interest disclosures

Dr. Dick is the Chief Scientific Officer of Thrive Genetics, Inc. Dr. Dick is also on the Advisory Board for the Seek Women’s Health Company. She owns stock in both companies. She received royalties from authoring the book, The Child Code: Understanding Your Child’s Unique Nature for Happier, More Effective Parenting, published by Avery, an imprint of the Penguin group. Dr Harvey reported receiving personal fees from Boehringer Ingelheim, Bioexcel, Karuna Therapeutics, Minerva Neuroscience, Alkermes, Sunovion, and Roche; royalties from WCG Endpoint Solutions; and equity from i-Function outside the submitted work. These did not impact or influence the collection, development, analysis, or interpretation of the contents of this manuscript. No other disclosures were reported.

## Data availability

All data sources are described in the manuscript and supplemental information. Only data from existing studies or study cohorts were analyzed, some of which have restricted access to protect the privacy of the study participants. The full summary statistics for the discovery MVP GWAS are available via dbGaP (accession: phs001672.v5.p1). Add Health genetic data can be obtained through dbGaP (accession: phs001367.v1.p1). COGA genetic data are available through dbGaP (accession: phs000763.v1.p1).

## Notes

### Competing Interest Statement

Dr. Dick is the Chief Scientific Officer of Thrive Genetics, Inc. Dr. Dick is also on the Advisory Board for the Seek Womens Health Company. She owns stock in both companies. She received royalties from authoring the book, The Child Code: Understanding Your Childs Unique Nature for Happier, More Effective Parenting, published by Avery, an imprint of the Penguin group. Dr Harvey reported receiving personal fees from Boehringer Ingelheim, Bioexcel, Karuna Therapeutics, Minerva Neuroscience, Alkermes, Sunovion, and Roche; royalties from WCG Endpoint Solutions; and equity from i-Function outside the submitted work. These did not impact or influence the collection, development, analysis, or interpretation of the contents of this manuscript. No other disclosures were reported.

### Author Declarations

All analyses were approved by the VA Central institutional review board.

### Summary of Updates

We have updated analyses and changed the focus of the discussion, highlighting the prospective analyses and moving other portions to the supplemental information.

## References

1. Case, A. & Deaton, A. Rising morbidity and mortality in midlife among white non-Hispanic Americans in the 21st century. Proc. Natl. Acad. Sci. U. S. A. 112, 15078–15083 (2015).

2. Tilstra, A. M., Simon, D. H. & Masters, R. K. Trends in ‘Deaths of Despair’ among Working-Aged White and Black Americans, 1990-2017. Am. J. Epidemiol. 190, 1751–1759 (2021).

3. Olfson, M. et al. National Trends in Suicide Attempts Among Adults in the United States. JAMA Psychiatry 74, 1095–1103 (2017).

4. Potash, J. B. et al. Attempted Suicide and Alcoholism in Bipolar Disorder: Clinical and Familial Relationships. Am J Psychiatry 157, 12 (2000).

5. Sher, L. Alcoholism and suicidal behavior: A clinical overview. Acta Psychiatrica Scandinavica vol. 113 13–22 Preprint at 10.1111/j.1600-0447.2005.00643.x (2006).

6. Koller, G., Preuß, U. W., Bottlender, M., Wenzel, K. & Soyka, M. Impulsivity and aggression as predictors of suicide attempts in alcoholics. Eur. Arch. Psychiatry Clin. Neurosci. 252, 155–160 (2002).

7. Whiteford, H. A. et al. Global burden of disease attributable to mental and substance use disorders: findings from the Global Burden of Disease Study 2010. The Lancet 382, 1575–1586 (2013).

8. Modesto-Lowe, V., Brooks, D. & Ghani, M. Alcohol dependence and suicidal behavior: From research to clinical challenges. Harvard Review of Psychiatry vol. 14 241–248 Preprint at 10.1080/10673220600975089 (2006).

9. Septier, M., Stordeur, C., Zhang, J., Delorme, R. & Cortese, S. Association between suicidal spectrum behaviors and Attention-Deficit/Hyperactivity Disorder: A systematic review and meta-analysis. Neurosci. Biobehav. Rev. 103, 109–118 (2019).

10. Pompili, M. et al. Epidemiology of suicide in bipolar disorders: a systematic review of the literature. Bipolar Disord. 15, 457–490 (2013).

11. Munafò, M. R., Tilling, K., Taylor, A. E., Evans, D. M. & Davey Smith, G. Collider scope: when selection bias can substantially influence observed associations. Int. J. Epidemiol. 47, 226–235 (2018).

12. Achenbach, T. M. The classification of children’s psychiatric symptoms: a factor-analytic study. Psychol. Monogr. 80, 1–37 (1966).

13. Kotov, R. et al. The Hierarchical Taxonomy of Psychopathology (HiTOP) in psychiatric practice and research. Psychol. Med. 52, 1666–1678 (2022).

14. Anttila, V. et al. Analysis of shared heritability in common disorders of the brain. Science (1979). 360, eaap8757 (2018).

15. Young, S. E., Stallings, M. C., Corley, R. P., Krauter, K. S. & Hewitt, J. K. Genetic and environmental influences on behavioral disinhibition. Am. J. Med. Genet. 96, 684–695 (2000).

16. Hicks, B. M., Schalet, B. D., Malone, S. M., Iacono, W. G. & McGue, M. Psychometric and Genetic Architecture of Substance Use Disorder and Behavioral Disinhibition Measures for Gene Association Studies. Behav. Genet. 41, 459–475 (2011).

17. Krueger, R. F. et al. Etiological connections among substance dependence, antisocial behavior and personality: Modeling the externalizing spectrum. J. Abnorm. Psychol. 111, 411–424 (2002).

18. Karlsson Linnér, R. et al. Multivariate analysis of 1.5 million people identifies genetic associations with traits related to self-regulation and addiction. Nat. Neurosci. 1–10 (2021) doi:10.1038/s41593-021-00908-3.

19. Baselmans, B. et al. The Genetic and Neural Substrates of Externalizing Behavior. Biological Psychiatry Global Open Science 2, 389–399 (2022).

20. Clapp Sullivan, M. L. et al. Beyond the factor indeterminacy problem using genome-wide association data. Nat. Hum. Behav. 8, 205–218 (2024).

21. Mullins, N. et al. Dissecting the Shared Genetic Architecture of Suicide Attempt, Psychiatric Disorders, and Known Risk Factors. Biol. Psychiatry 91, 313–327 (2022).

22. Bigdeli, T. B. et al. Correlates of suicidal behaviors and genetic risk among United States veterans with schizophrenia or bipolar I disorder. Mol. Psychiatry 29, 2399–2407 (2024).

23. Barr, P. B. et al. Correlates of Risk for Disinhibited Behaviors in the Million Veteran Program Cohort. JAMA Psychiatry 81, 188–197 (2024).

24. Docherty, A. R. et al. Genome-Wide Association Study of Suicide Death and Polygenic Prediction of Clinical Antecedents. American Journal of Psychiatry 177, 917–927 (2020).

25. Yuval, B.-C. et al. Predicting Suicidal Behavior From Longitudinal Electronic Health Records. American Journal of Psychiatry 174, 154–162 (2017).

26. Edwards, A. C. et al. Alcohol use disorder and non-fatal suicide attempt: findings from a Swedish National Cohort Study. Addiction 117, 96–105 (2022).

27. Gaziano, J. M. et al. Million Veteran Program: A mega-biobank to study genetic influences on health and disease. J. Clin. Epidemiol. 70, 214–223 (2016).

28. Kramarow, E. A. & Pastor, P. N. The health of male veterans and nonveterans aged 25-64: United States, 2007-2010. NCHS Data Brief 1–8 (2012).

29. Boersma, P., Cohen, R. A., Zelaya, C. E. & Moy, E. Multiple Chronic Conditions Among Veterans and Nonveterans: United States, 2015-2018. Natl. Health Stat. Report. 1–13 (2021).

30. Hunter-Zinck, H. et al. Genotyping Array Design and Data Quality Control in the Million Veteran Program. Am. J. Hum. Genet. 106, 535–548 (2020).

31. Fang, H. et al. Harmonizing Genetic Ancestry and Self-identified Race/Ethnicity in Genome-wide Association Studies. Am. J. Hum. Genet. 105, 763–772 (2019).

32. Taliun, D. et al. Sequencing of 53,831 diverse genomes from the NHLBI TOPMed Program. Nature 590, 290–299 (2021).

33. Denny, J. C. et al. Systematic comparison of phenome-wide association study of electronic medical record data and genome-wide association study data. Nat. Biotechnol. 31, 1102–1110 (2013).

34. Wu, P. et al. Mapping ICD-10 and ICD-10-CM codes to phecodes: Workflow development and initial evaluation. J. Med. Internet Res. 21, 1–13 (2019).

35. Zheutlin, A. B. et al. Penetrance and pleiotropy of polygenic risk scores for schizophrenia in 106,160 patients across four health care systems. American Journal of Psychiatry 176, 846–855 (2019).

36. Bigdeli, T. B. et al. Penetrance and Pleiotropy of Polygenic Risk Scores for Schizophrenia, Bipolar Disorder, and Depression among Adults in the US Veterans Affairs Health Care System. JAMA Psychiatry 79, 1092–1101 (2022).

37. Zhou, W. et al. Efficiently controlling for case-control imbalance and sample relatedness in large-scale genetic association studies. Nat. Genet. 212357 (2018) doi:10.1038/s41588-018-0184-y.

38. Grotzinger, A. D. et al. Genomic structural equation modelling provides insights into the multivariate genetic architecture of complex traits. Nat. Hum. Behav. 3, 513–525 (2019).

39. Lee, P. H. et al. Genomic relationships, novel loci, and pleiotropic mechanisms across eight psychiatric disorders. Cell 179, 1469–1482 (2019).

40. Waldman, I. D., Poore, H. E., Luningham, J. M. & Yang, J. Testing structural models of psychopathology at the genomic level. World Psychiatry 10.1002/wps.20772 (2020) doi:10.1002/wps.20772.

41. Luo, Y. et al. Estimating heritability and its enrichment in tissue-specific gene sets in admixed populations. Hum. Mol. Genet. 30, 1521–1534 (2021).

42. Watanabe, K., Taskesen, E., van Bochoven, A. & Posthuma, D. Functional mapping and annotation of genetic associations with FUMA. Nat. Commun. 8, 1–11 (2017).

43. Ashley-Koch, A. E. et al. Genome-wide association study identifies four pan-ancestry loci for suicidal ideation in the Million Veterans Program. PLoS Genet. 19, e1010623 (2023).

44. Frei, O. et al. Bivariate causal mixture model quantifies polygenic overlap between complex traits beyond genetic correlation. Nat. Commun. 10, 2417 (2019).

45. Agrawal, A. et al. The Collaborative Study on the Genetics of Alcoholism: Overview. Genes Brain Behav. 22, e12864 (2023).

46. Begleiter, H. The Collaborative Study on the Genetics of Alcoholism. Alcohol Health Res. World 19, 228–236 (1995).

47. Dick, D. M. et al. The collaborative study on the genetics of alcoholism: Sample and clinical data. Genes Brain Behav. 22, e12860 (2023).

48. Bucholz, K. K. et al. A new, semi-structured psychiatric interview for use in genetic linkage studies: a report on the reliability of the SSAGA. J. Stud. Alcohol 55, 149–58 (1994).

49. Johnson, E. C. et al. The collaborative study on the genetics of alcoholism: Genetics. Genes Brain Behav. 22, e12856 (2023).

50. Harris, K. M. et al. The National Longitudinal Study of Adolescent to Adult Health: research design. Add Health: The National Longitudinal Study of Adolescent to Adult Health Preprint at http://www.cpc.unc.edu/projects/addhealth/design (2009).

51. Ruan, Y. et al. Improving polygenic prediction in ancestrally diverse populations. Nat. Genet. 54, 573–580 (2022).

52. Ding, Y. et al. Polygenic scoring accuracy varies across the genetic ancestry continuum. Nature 618, 774–781 (2023).

53. Putter, H., Fiocco, M. & Geskus, R. B. Tutorial in biostatistics: competing risks and multi-state models. Stat. Med. 26, 2389–2430 (2007).

54. Fine, J. P. & Gray, R. J. A Proportional Hazards Model for the Subdistribution of a Competing Risk. J. Am. Stat. Assoc. 94, 496–509 (1999).

55. Terry M. Therneau & Patricia M. Grambsch. Modeling Survival Data: Extending the Cox Model. (Springer, New York, 2000).

56. Therneau, T. M. A Package for Survival Analysis in R. Preprint at https://CRAN.R-project.org/package=survival (2024).

57. Blosnich, J. R. et al. Social Determinants and Military Veterans’ Suicide Ideation and Attempt: a Cross-sectional Analysis of Electronic Health Record Data. J. Gen. Intern. Med. 35, 1759–1767 (2020).

58. Coimbra, B. M. et al. Meta-analysis of the effect of racial discrimination on suicidality. SSM Popul. Health 20, 101283 (2022).

59. Paradies, Y. et al. Racism as a Determinant of Health: A Systematic Review and Meta-Analysis. PLoS One 10, e0138511- (2015).

60. Peterson, R. E. et al. Genome-wide Association Studies in Ancestrally Diverse Populations: Opportunities, Methods, Pitfalls, and Recommendations. Cell 179, 589–603 (2019).

61. Willer, C. J., Li, Y. & Abecasis, G. R. METAL: fast and efficient meta-analysis of genomewide association scans. Bioinformatics 26, 2190–2191 (2010).

62. Docherty, A. R. et al. GWAS Meta-Analysis of Suicide Attempt: Identification of 12 Genome-Wide Significant Loci and Implication of Genetic Risks for Specific Health Factors. American Journal of Psychiatry 180, 723–738 (2023).

63. Adams, M. J. et al. Trans-ancestry genome-wide study of depression identifies 697 associations implicating cell types and pharmacotherapies. Cell 188, 640–652.e9 (2025).

64. Trubetskoy, V. et al. Mapping genomic loci implicates genes and synaptic biology in schizophrenia. Nature 604, 502–508 (2022).

65. Edwards, A. C. et al. The role of substance use disorders in the transition from suicide attempt to suicide death: a record linkage study of a Swedish cohort. Psychol. Med. 54, 90–97 (2024).

66. Ferrari, A. J. et al. The burden attributable to mental and substance use disorders as risk factors for suicide: Findings from the Global Burden of Disease Study 2010. PLoS One 9, e91936 (2014).

67. Lannoy, S. et al. Risk of suicidal behavior as a function of alcohol use disorder typologies: A Swedish population-based study. Addiction 119, 281–290 (2024).

68. Colbert, S. M. C. et al. Exploring the genetic overlap of suicide-related behaviors and substance use disorders. American Journal of Medical Genetics, Part B: Neuropsychiatric Genetics 186, 445–455 (2021).

69. Colbert, S. M. C. et al. Polygenic Contributions to Suicidal Thoughts and Behaviors in a Sample Ascertained for Alcohol Use Disorders. Complex Psychiatry 9, 11–23 (2023).

70. Yan, C. et al. Differences in Health Professionals’ Engagement With Electronic Health Records Based on Inpatient Race and Ethnicity. JAMA Netw. Open 6, e2336383–e2336383 (2023).

71. Boardman, J. D., Daw, J. & Freese, J. Defining the environment in gene-environment research: Lessons from social epidemiology. Am. J. Public Health 103, S64--S72 (2013).

72. Barr, P. B. et al. Clinical, environmental, and genetic risk factors for substance use disorders: characterizing combined effects across multiple cohorts. Mol. Psychiatry 27, 4633–4641 (2022).

