## Supplemental info for "The Externalizing Spectrum and Suicide Risk: Insights from Genomics and Electronic Health Records in over 500,000 Veterans"

### VA Million Veteran Program Core Acknowledgements for Publications

**JUNE 2025**

**MVP PROGRAM OFFICE**

- Sumitra Muralidhar, Ph.D., Program Director

US Department of Veterans Affairs, 810 Vermont Avenue NW, Washington, DC 20420

- Jennifer Moser, Ph.D., Associate Director, Scientific Programs

US Department of Veterans Affairs, 810 Vermont Avenue NW, Washington, DC 20420

- Jennifer E. Deen, B.S., Associate Director, Cohort & Public Relations

US Department of Veterans Affairs, 810 Vermont Avenue NW, Washington, DC 20420

**MVP EXECUTIVE COMMITTEE**

- Co-Chair: Philip S. Tsao, Ph.D.

VA Palo Alto Health Care System, 3801 Miranda Avenue, Palo Alto, CA 94304

- Co-Chair: Sumitra Muralidhar, Ph.D.

US Department of Veterans Affairs, 810 Vermont Avenue NW, Washington, DC 20420

- J. Michael Gaziano, M.D., M.P.H.

VA Boston Healthcare System, 150 S. Huntington Avenue, Boston, MA 02130

- Elizabeth Hauser, Ph.D.

Durham VA Medical Center, 508 Fulton Street, Durham, NC 27705

**-**	Amy Kilbourne, Ph.D., M.P.H.

VA HSR&D, 2215 Fuller Road, Ann Arbor, MI 48105

- Michael Matheny, M.D., M.S., M.P.H.

VA Tennessee Valley Healthcare System, 1310 24th Ave. South, Nashville, TN 37212

- Dave Oslin, M.D.

Philadelphia VA Medical Center, 3900 Woodland Avenue, Philadelphia, PA 19104

- Deepak Voora, MD

Durham VA Medical Center, 508 Fulton Street, Durham, NC 27705

**MVP CO-PRINCIPAL INVESTIGATORS**

- J. Michael Gaziano, M.D., M.P.H.

VA Boston Healthcare System, 150 S. Huntington Avenue, Boston, MA 02130

- Philip S. Tsao, Ph.D.

VA Palo Alto Health Care System, 3801 Miranda Avenue, Palo Alto, CA 94304

**MVP CORE OPERATIONS**

- Jessica V. Brewer, M.P.H., Director, MVP Cohort Operations

VA Boston Healthcare System, 150 S. Huntington Avenue, Boston, MA 02130

- Mary T. Brophy M.D., M.P.H., Director, VA Central Biorepository

VA Boston Healthcare System, 150 S. Huntington Avenue, Boston, MA 02130

- Kelly Cho, M.P.H, Ph.D., Director, MVP Phenomics

VA Boston Healthcare System, 150 S. Huntington Avenue, Boston, MA 02130

**-**	Lori Churby, B.S., Director, MVP Regulatory Affairs

VA Palo Alto Health Care System, 3801 Miranda Avenue, Palo Alto, CA 94304

- Scott L. DuVall, Ph.D., Director, VA Informatics and Computing Infrastructure (VINCI) VA Salt Lake City Health Care System, 500 Foothill Drive, Salt Lake City, UT 84148
- Saiju Pyarajan Ph.D., Director, Data and Computational Sciences

VA Boston Healthcare System, 150 S. Huntington Avenue, Boston, MA 02130

- Robert Ringer, Pharm.D., Director, VA Albuquerque Central Biorepository

New Mexico VA Health Care System, 1501 San Pedro Drive SE, Albuquerque, NM 87108

- Luis E. Selva, Ph.D., Director, MVP Biorepository Coordination

VA Boston Healthcare System, 150 S. Huntington Avenue, Boston, MA 02130

- Shahpoor (Alex) Shayan, M.S., Director, MVP PRE Informatics

VA Boston Healthcare System, 150 S. Huntington Avenue, Boston, MA 02130

- Brady Stephens, M.S., Principal Investigator, MVP Information Center Canandaigua VA Medical Center, 400 Fort Hill Avenue, Canandaigua, NY 14424
- Stacey B. Whitbourne, Ph.D., Director, MVP Cohort Development and Management VA Boston Healthcare System, 150 S. Huntington Avenue, Boston, MA 02130

### Univariate Genome Wide Association Studies (GWAS)

Outcomes for the univariate GWASs included in the subsequent multivariate models came from electronic health records (EHR), the MVP Baseline Survey, and the MVP Lifestyle survey. EHR data were phecodes, which are clusters of ICD-9/10-CM codes ^1,2^. We considered individuals as having a lifetime diagnosis for any given phecode if there were two or more occurrences of that phecode in their EHR, consistent with prior EHR analyses ^3,4^. We used phecodes for *Substance addiction and disorders* (Phecode 316, DUD), *Alcohol-related disorders* (Phecode 317, AUD), *Tobacco use disorder* (Phecode 318, TUD), and *Attention deficit hyperactivity disorder* (Phecode 313.1, ADHD). From the Baseline surveys we used “In your lifetime, have you smoked a total of at least 100 cigarettes, cigars, or pipes?” for lifetime smokers (SMOK), and “How often do you have six or more drinks on one occasion?” for binge drinking (BINGE).

We performed all univariate GWASs using SAIGE ^5^, to adjust for relatedness. In Step 1 of SAIGE we filtered down to a set of 77,215 LD independent (*r^2^* < 0.1), genotyped single nucleotide polymorphisms (SNPs) in the EUR-like veterans and 147,496 independent, genotyped SNPs in the AFR-like veterans using a 50kb window and a 10bp step size. These SNPs were used to create the genetic relatedness matrices (GRM) used in Step 2. Next (Step 2), we performed association tests using a leave-one-chromosome-out (LOCO) approach so that the chromosome which included the variant being tested was excluded from the calculation of the GRM. Including the chromosome which has the SNP being tested in the GRM can lead to a deflation in heritability estimates and unnecessarily reduce power ^6^ for follow up in GenomicSEM. All analyses included age, sex, and the first 20 genetic principal components as covariates. The descriptive statistics and GWAS metrics for each of the GWASs from SAIGE are presented for each population in the table below. With the exception of ADHD in the AFR-like veterans, all of the GWASs had significant SNP-based heritability.

| **SAIGE Results for GWASs used in GenomicSEM** | | | | | | | |
| --- | --- | --- | --- | --- | --- | --- | --- |
| Label | Population | Cases | Controls | *h^2^*_SNP_ (SE) | LDSC int. | Mean χ^2^ | λ_GC_ |
| ADHD | EUR-like | 11,154 | 428,978 | 0.073 (0.013) | 1.014 | 1.076 | 1.071 |
| AUD | EUR-like | 76,162 | 344,390 | 0.101 (0.005) | 1.024 | 1.529 | 1.414 |
| DUD | EUR-like | 49,751 | 373,936 | 0.113 (0.005) | 1.036 | 1.435 | 1.350 |
| TUD | EUR-like | 142,419 | 255,162 | 0.091 (0.003) | 1.065 | 1.749 | 1.570 |
| SMOK | EUR-like | 223,696 | 82,705 | 0.114 (0.005) | 1.009 | 1.559 | 1.424 |
| BINGE | EUR-like | 191,545 | - | 0.053 (0.004) | 1.035 | 1.238 | 1.216 |
| ADHD | AFR-Like | 946 | 113,835 | 0.096 (0.070) | 0.999 | 1.011 | 1.008 |
| AUD | AFR-Like | 34,601 | 73,933 | 0.050 (0.006) | 1.008 | 1.154 | 1.139 |
| DUD | AFR-Like | 30,467 | 78,772 | 0.049 (0.006) | 1.016 | 1.150 | 1.140 |
| TUD | AFR-Like | 43,712 | 61,705 | 0.049 (0.005) | 1.010 | 1.172 | 1.155 |
| SMOK | AFR-Like | 33,816 | 17,219 | 0.050 (0.008) | 0.998 | 1.071 | 1.061 |
| BINGE | AFR-Like | 32,627 | - | 0.055 (0.011) | 1.030 | 1.088 | 1.094 |
| MOS-ATTN | AFR-Like | 33,960 | - | 0.033 (0.009) | 1.003 | 1.038 | 1.035 |

### multivariate genome wide association study (GWAS)

We fit initial models with 6 indicators (AUD, DUD, TUD, ADHD, SMOK, and BINGE). Our final model for the EUR-like analyses contained all six indicators, with correlated residuals between the binge drinking and AUD and lifetime smoking and TUD indicators, respectively. The data showed reasonably good fit to this model specification (Figure 1, Panel A; *χ^2^*= 104.10, df = 7, p = 1.53x10^-19^; CFI = 0.99, SRMR = 0.06). For the AFR-like veterans, the genetic correlation between SMOK and TUD was indistinguishable from one, so we excluded SMOK and retained TUD as it had greater statistical power. We also excluded ADHD due to the low power of the GWAS, replacing it with the attention problems subscale of the Medical Outcomes Survey (MOS-ATTN, *h*^2^_SNP_ [SE] = 0.033 [0.009], LDSC intercept = 1.003, Mean χ^2^ = 1.038, λ_GC_ = 1.035), which correlated with ADHD ~ 0.8, as a proxy. The final model for AFR-like veterans therefore, largely mimicked that of the EUR-like veterans with one fewer indicator (Figure 1, Panel B; *χ^2^*= 131.51, df = 8, p = 1.38x10^-24^; CFI = 0.96, SRMR = 0.09).

After QC, we used the final models for each population to perform a multivariate GWAS across 8,142,519 SNPs in the EUR-like and 14,101,162 SNPs in the AFR-like populations. We identified 134 and 1 genome wide significant loci in EUR-like and AFR-like populations, respectively. We then performed a meta-analysis using an inverse-variance weighted fixed effects meta-analysis in METAL ^7^. There were 155 independent genome wide significant loci in the meta-analysis. Supplmemntary Figure 1 presents the Miami plot for the meta-analysis results, with the EUR-like Q-SNP results on the left. Importantly, only 11 of the 155 loci were significant using a conservative threshold in the heterogeneity tests (*p* <.05/155 = 3.23 x10^-4^), suggesting that many of these SNPs are associated through the latent factor. Of the 138 loci present in the Externalizing Consortium GWAS (EXT1.0) ^8^ which did not include any MVP data, 94.2% (95% CI = 88.9%, 97.5%) were sign concordant and, 116 (84.1%) of the loci were at least nominally significant (p < .05) in EXT1.0. Correspondingly, of the 567 of 579 loci from EXT1.0 present in the current GWAS, 95.6% are in the same direction and 388 (68.4%) are nominally significant. Full results for the meta-analysis and each population are listed in Supplemental Tables 2 and 3.

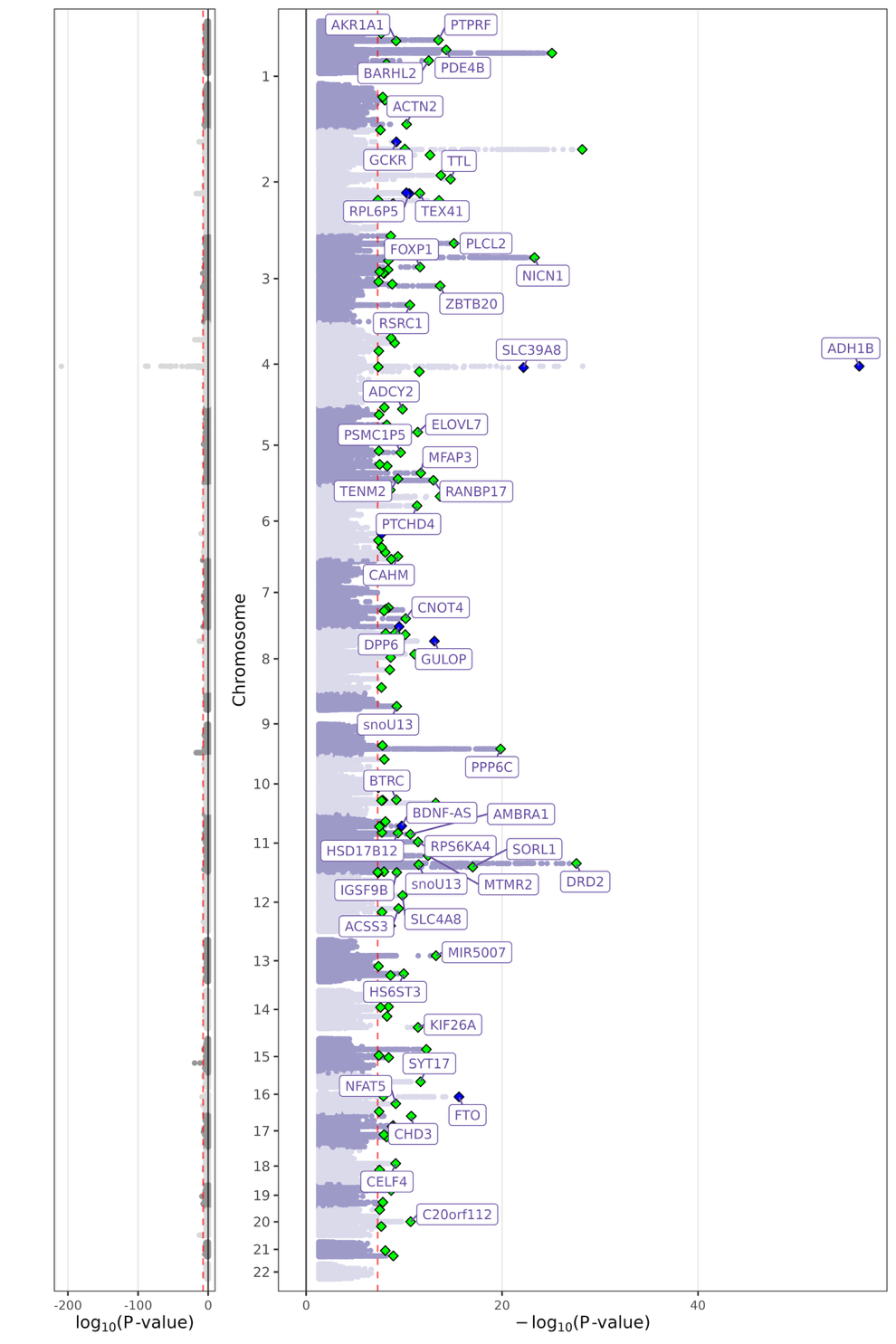

Supplemental Figure 1 Figure 1: Trans-ancestry meta-analysis results for externalizing in MVP.

Miami plot for trans-ancestry meta-analysis (Q-SNP results on the left, multivariate GWAS on the right). Genome wide significant loci indicated by colored diamonds. Green diamonds = loci not significant in Q-SNP analyses. Blue diamonds = loci that are also Q-SNPs.

#### Validating MVP-EXT using external results and genetic correlations

Next, in order to ensure that our latent externalizing factor in MVP was capturing broad externalizing risk and not just risk for SUD, we conducted a series of comparisons between the current externalizing factor (MVP-EXT) and the results from the Externalizing Consortium’s GWAS (EXT1.0) First, we estimated the genetic correlation between the two latent factors in Genomic SEM, which demonstrated a very strong overlap (rG = 0.87, 95% CI = 0.83, 0.91). Second, we compared the genetic correlations of MVP-EXT and EXT1.0 with 93 external traits (Supplemental Figure 2, Panel A). Overall, the correlation of rG estimates was very strong (*r* = 0.96, 95% CI = 0.93, 0.97). Supplemental Figure 2, Panel B displays a selection of rG estimates that had the absolute largest discrepancies between MVP-EXT and EXT1.0. The results for MVP-EXT generally showed stronger associations with substance-related phenotypes. The one result where EXT1.0 was stronger was with risky behaviors. Overall, these results seem to suggest that MVP-EXT is largely recapitulating results from EXT1.0, even though the configuration of input phenotypes varies slightly between the two analyses (see Supplemental Table 4 for specific results).

Lastly, we compared genetic correlation estimates across the EUR-like and AFR-like results with external traits that had population-matched results. Panel C of Supplemental Figure 2 presents results side by side. Overall, the genetic correlations were consistent across populations with two exceptions. The genetic correlation between MVP-EXT and AUD was significantly stronger EUR-like veterans (rG = 0.96, 95% CI = 0.92, 0.99) compared to AFR-like veterans (rG = 0.79, 95% CI = 0.67, 0.91) and the genetic correlation between MVP-EXT and SCZ was significantly stronger AFR-like veterans (rG = 0.54, 95% CI = 0.42, 0.66) compared to EUR-like veterans (rG = 0.32, 95% CI = 0.28, 0.36). It is worth noting these differences are not large and could be largely influenced bias in diagnoses and interactions with the healthcare system, which are well documented for both SUD and schizophrenia ^9–11^. Genetic correlations between MVP-EXT and suicide attempt (SA) were comparable for both the EUR-like (rG = 0.67, 95% CI = 0.60, 0.91) and AFR-like (rG = 0.74, 95% CI = 0.42, 0.81) veterans. Genetic correlations with SI were weak, but significant the EUR-like (rG = 0.12, 95% CI = 0.08, 0.17) veterans and null for AFR-like (rG = -0.01, 95% CI = -0.20, 0.18) veterans. However, while the rG’s with SI were significant in EUR-like but not AFR-like participants, the confidence intervals were overlapping suggesting the lack of association in AFR-like veterans may reflect the lower power in the GWAS results.

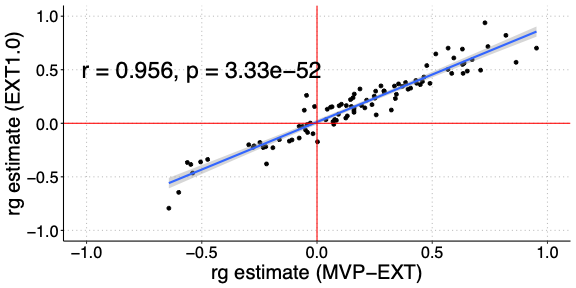

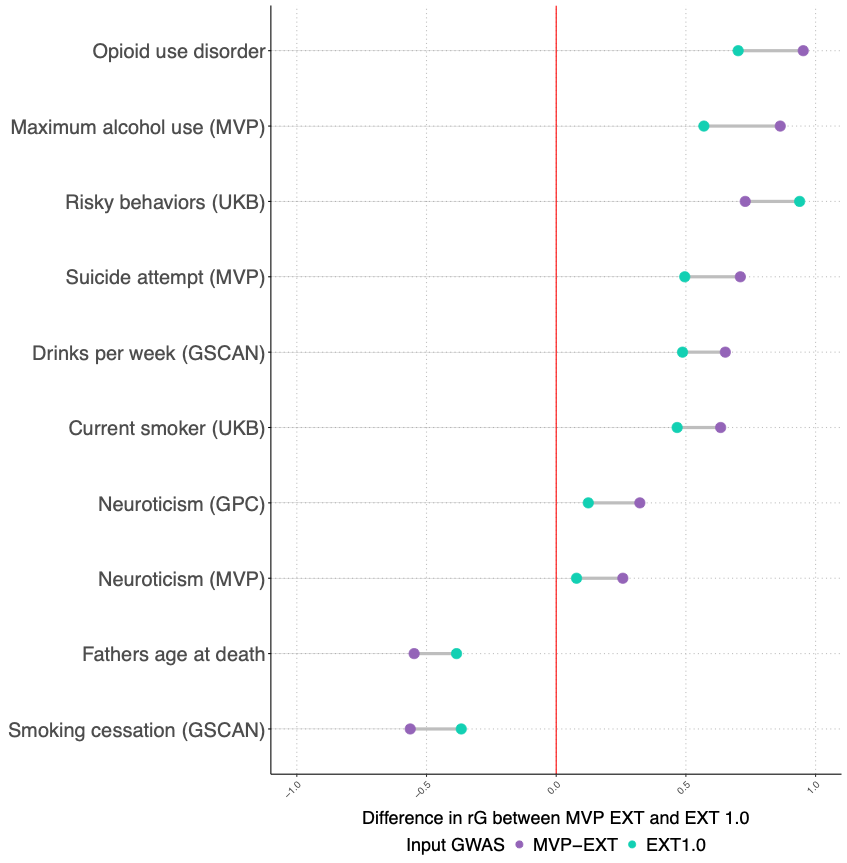

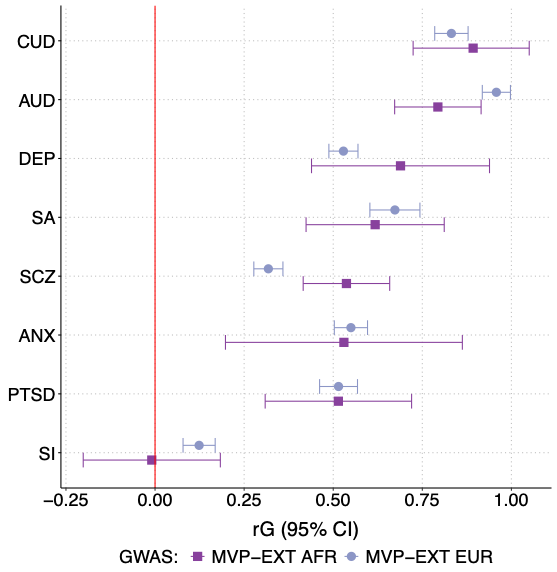

A

B

C

Supplemental Figure 2: Genetic correlations across EXT results and populations

(A) Correlation of genetic correlations between MVP-EXT and EXT1.0 across 93 external traits. (B) Top 10 genetic correlation estimates with the absolute largest discrepancy between MVP-EXT and EXT1.0 (EUR-like only). (3) Comparison of MVP-EXT and external traits across EUR-like and AFR-like results.

### Post GWAS pipeline

Our post-GWAS, bioinformatic pipeline had several steps. First, we tested gene-level and gene-set enrichment analysis using MAGMA (version 1.08)^12^, and its recent intersessions (FUMA version 1.3.6)^13^. In all the MAGMA-based analyses, genome-wide SNPs were mapped to 18,235 protein-coding genes from Ensembl v102, with a 1 kb window for both sides (i.e., start and end). We corrected all MAGMA-based associations for multiple-testing using a Bonferroni correction (one-sided *p* < 2.74x10^-6^). Second, we used MetaXcan^20^ to conduct a Transcriptome-Wide Association Study (TWAS) using genetically regulated expression models from GTEx v8^42^. We applied within-tissue FDR correction identify significant TWAS associations. Finally, we used summary-data-based Mendelian randomization (SMR) ^21^ to whether gene expression mediated the relationship between SNPs and the phenotype and distinguish causally-related and pleiotropic models using the heterogeneity in dependent instruments (HEIDI) test. The latter test was used to identify genes that are more likely to be functionally relevant to the phenotype and should therefore be prioritized for follow up. We identified genes of interest as those that met SMR test Bonferroni correction significance threshold and had a HEIDI test p-value > .05.

We applied the single-cell Disease-Relevance Scoring (scDRS)^14^ method to assess the collective expression of candidate disease-associated genes derived from EUR-like summary statistics using MAGMA. Each potential disease gene was weighted by its MAGMA Z-score from the GWAS results and adjusted for gene-specific technical noise in single-cell data obtained from an extensive postmortem dataset, consisting of 450K cells from the dorsolateral prefrontal cortex (DLPFC) of 40 human donors from the VA’s National PTSD Brain Bank (NPBB) ^15^ , described in detail elsewhere ^16^. We conducted snRNA-seq analysis on DLPFC samples from 16 “control” individuals (non-suicides related death) and 24 “cases” (confirmed suicide deaths), which were further divided into post-traumatic stress disorder (PTSD, n=12) and major depressive disorder (MDD, n=12) subgroups. The scDRS approach produced cell-specific raw disease scores. To ensure rigor, we generated 1,000 sets of cell-specific raw control scores using matched control gene sets, ensuring consistency in gene set size, mean expression, and expression variance with the candidate disease genes. We then normalized both the raw disease and control scores for each cell, resulting in normalized disease and control scores. These calculations were performed using the default parameters of the scDRS compute-score function. For further analysis, we evaluated cell type-level associations to identify broad cell types linked to the disease and to examine variability in disease association across individual cells within each cell type. This analysis was conducted using the scDRS perform-downstream() function with default settings. To account for multiple testing, we applied the Benjamini-Hochberg method to calculate the false discovery rate (FDR)^17^.

#### Results

Counts of significant gene-based associations from MAGMA, TWAS, and SMR are presented in Supplemental Figure 3A (238, 372, and 181, respectively). While many of the associations appear to be method specific, there is considerable overlap, with 172 of these total associations being detected in more than one method (full results available in Supplemental Tables 5 – 7).

Lastly, we examined whether the genetic architecture of externalizing converges to similar cell type enrichment in a sample composed of both suicide deaths and deaths from other causes (Supplemental Figure 4A). Supplemental Figure 4B demonstrates the cell-type clusters for comparison. We observed significant enrichment within inhibitory neurons, astroglia, and oligodendrocyte progenitor cells (OPCs) in the meta-analyzed results (Supplemental Figure 4C); however, only the inhibitory neurons were significant in the AFR-like results, a finding that is likely attributable to lower statistical power. Recent analyses in other psychiatric conditions, including schizophrenia ^18^ and PTSD ^16^ have found evidence of enrichment in these neurons. Interestingly, while others have also found evidence of excitatory neurons for these psychiatric conditions ^16,18^ and suicide attempt ^19^, they were not relevant externalizing in a sample comprised primarily of suicide deaths. The differences between our results and prior analyses could reflect distinct biological pathways, but could also reflect the limited power in our post-mortem brain samples (N = 40). Results from these analyses should be interpreted cautiously until they can be replicated in larger cohorts.

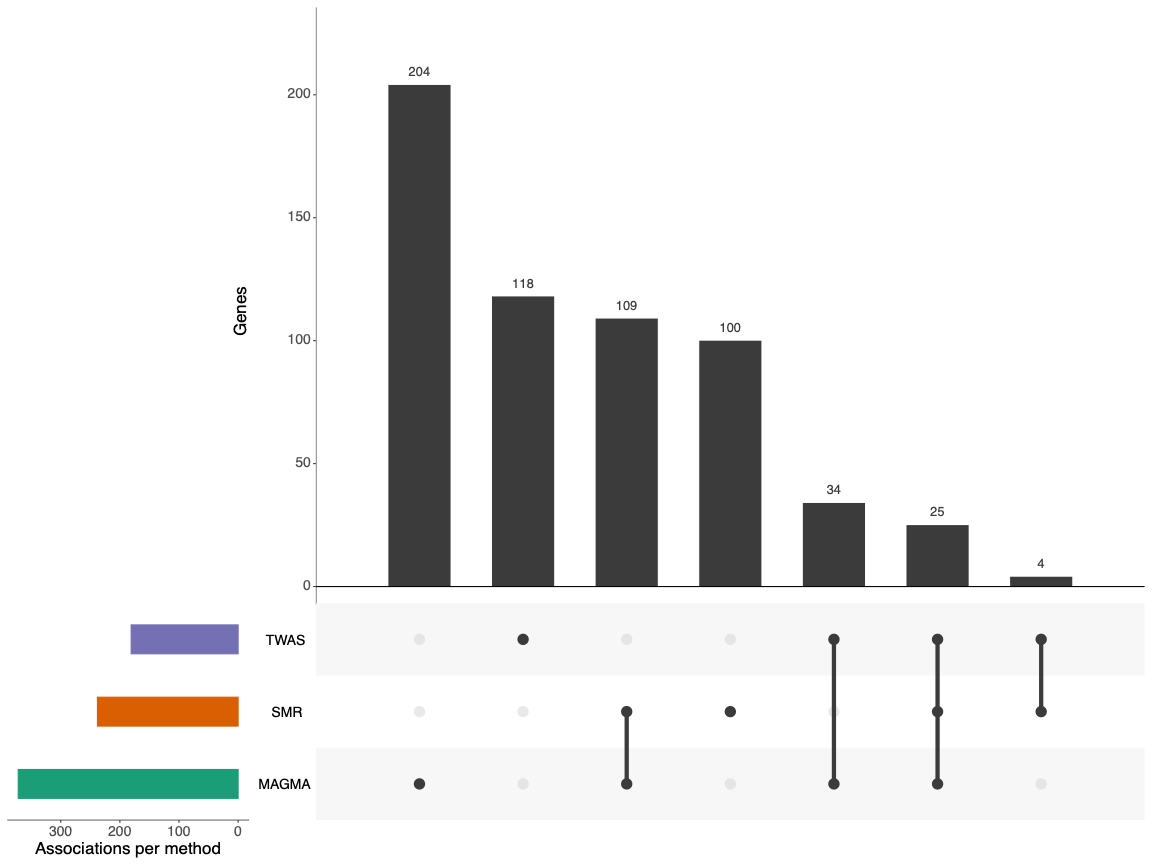

Supplemental Figure 3: Overlap in results from post-GWAS MVP-EXT analyses

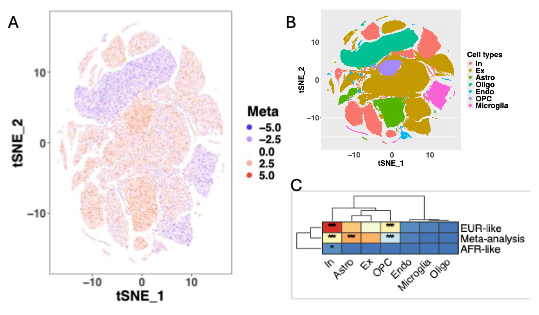

Supplemental Figure 4: Cell-type enrichment for MVP-EXT results in post-mortem brain tissue.

Cell type enrichment across suicide deaths and deaths from other causes. Scale is in Z-scores (Panel A). Cell-type clusters (Panel B). Tests for cell-type enrichment across European-like, African-like, and meta-analysis results (Panel C).

### External validation samples

#### The Collaborative Study on the Genetics of Alcoholism (COGA)

COGA is a multi-site study of families affected with AUD, designed to identify and understand genetic factors involved in the predisposition to alcohol and other substance use disorders ^20–22^. Probands and first-degree relatives were assessed, with recruitment extended to include additional relatives and community ascertained comparison families (N = 17,878). Participants completed a poly-diagnostic interview, the Semi-Structured Assessment for the Genetics of Alcoholism (SSAGA) ^23,24^. We currently have genome wide data on 12,145 individuals. Genetic data were used to assign individuals into genetically similar groupings ^25^ based on the first two principal components and the 1000 genomes reference panel (Phase 3, version 5) ^26^. Families were classified as primarily European-like (EUR-like) or African-like (AFR-like) according to the genetic similarity of the greatest proportion of family members ^27^. Full details of genotyping, imputation and quality control have been described elsewhere ^26,27^. The final analytic sample consisted of 10,986 COGA participants (N_EUR-like_ = 7,601; N_AFR-like_ = 3,385).

#### The National Longitudinal Study of Adolescent to Adult Health (Add Health)

Add Health is an ongoing, nationally representative longitudinal study participants recruited as adolescents (ages 11-18) and followed into adulthood (ages 35-42) in the United States ^28^. Add Health participants were selected from a stratified sample of 132 schools resulting in an initial, nationally representative sample of 90,118 students in grades 7-12. Of the original sample, 20,745 were selected for additional in-home interviews. In total, 15,159 individuals interviewed during Wave IV (ages 24-32) provided samples for genotyping, conducted using the Illumina Omni1 and Omni2.5 arrays. After quality control, genotypic data are available for 9,974 individuals. Genotypes for those assigned as most similar to European reference panels were imputed to the Haplotype Reference Consortium (HRC) reference panel. Data for all other populations were imputed to the 1000 Genomes, Phase III reference panel. Our final analytic sample consisted of 6,883 individuals (N_EUR-like_ = 5,122; N_AFR-like_ = 1,761).

#### Phenotype definitions

##### Externalizing factor in COGA and Add Health

The phenotypes included for the latent externalizing factor in both Add Health and COGA matched the indicators from the multivariate GWAS model, one-to-one. Both Add Health and COGA contained some form of clinical interview. In Add Health, respondents received the Composite International Diagnostic Interview-Substance Abuse Module (CIDI-SAM) ^29^ at Wave IV (ages 24-32). In COGA, respondents received the Semi-Structured Assessment for the Genetics of Alcoholism (SSAGA) ^23,24^ at all assessments.

Attention deficit hyperactivity disorder (ADHD): In Add Health, we measured ADHD using a retrospective scale containing 18 items with responses ranging from “never or rarely” (0) to “very often” (3) and an overall scale ranging from 0 to 54. *ADHD* in COGA is measured using DSM-III-R/IV ADHD symptom counts from the SSAGA ^23,24^.

Alcohol use disorder (AUD): In Add Health, *alcohol use disorder (AUD)* was measured from the combined criteria counts of DSM-IV alcohol dependence and abuse (range 0 to 11) at Wave IV. In COGA, *alcohol use disorder* was measured from criteria counts of DSM-5 alcohol use disorder symptoms (range 0 to 11). The only differences between combining abuse and dependence from DSM-IV criteria and using the DSM-5 criteria (which mostly reflects the combination of abuse and dependence into a single disorder) are in a single item. DSM-IV abuse contains the criteria of “[i]n the past year, have you more than once gotten arrested, been held at a police station, or had other legal problems because of your drinking?” which was not included in DSM-5. Instead, DSM-5 added, “[i]n the past year, have you wanted to drink so badly you couldn’t think of anything else.”

Drug use disorder (DUD): In Add Health, *drug use disorder (AUD)* was measured from the combined criteria counts of DSM-IV dependence and abuse for cannabis or other substances (sedatives, tranquilizers, stimulants, painkillers, steroids, cocaine, crystal meth, and/or some other illicit substance), selecting whichever was greater (range 0 to 11). In COGA, *drug use disorder* was measured from the maximum criteria count of DSM-5 substance use disorder symptoms (cannabis, opioid, cocaine, stimulants, or sedatives; range 0 to 11). Again, the only differences between combined DSM-IV abuse and dependence and DSM-5 criteria is the cravings item.

Tobacco use disorder (TUD): In both Add Health and COGA, we used the Fagerstrom test for nicotine dependence (FTND) to assess *tobacco use disorders*. The FTND assesses six criteria and has values ranging from 0 to 10.

Lifetime smoking initiation (SMOK) In Add Health, SMOK was constructed as a binary measure from the question “Have you ever smoked cigarettes regularly, that is, at least 1 cigarette every day for 30 days?” If individuals indicated yes at any point in the four waves of data, they were coded as being a smoker. Individuals who answered no across all waves were coded as never being a smoker. In COGA, individuals were classified as yes on SMOK if they ever answered yes to “Over your lifetime, have you smoked a total of 100 cigarettes (smoked 5 or more packs)?” Individuals who answered no on the initial interview or across each point of data collection (for those who were interviewed more than once) were coded as never being a smoker.

Binge drinking frequency (BINGE): In Add Health, binge drinking was the lifetime maximum response to the question: “Over the past 12 months, on how many days did you drink 5 or more drinks in a row?” Responses ranged from 0 = “Never” to 6 = “Every day or almost every day”. In COGA, respondents were asked: “Now I'd like you to think about the last 12 months. How often did you have 5 or more drinks in a 24-hour period?” Responses ranged from 0 = “Never” to 12 = “Every day”.

##### Suicide-related outcomes

Suicidal ideation: In Add Health, suicidal ideation was assessed for the past year by asking respondents: “During the past 12 months, did you ever seriously think about committing suicide?” In COGA, respondents were asked about lifetime suicidal ideation at each assessment (“Have you ever thought about killing yourself?”). For both cohorts, individuals were coded as having suicidal ideation if they answered “Yes” at any point during data collection.

Suicide attempt: For suicide attempt, Add Health participants were asked “During the past 12 months, how many times did you actually attempt suicide?”, but only amongst those who indicated “Yes” for suicidal ideation. In COGA, respondents were asked: “Have you ever tried to kill yourself?” at each assessment. As in the case of ideation, participants were coded as having a lifetime suicide attempt if they answered more than 0 number of attempts (Add Health) or “Yes” (COGA) at any point.

### References

1.	Denny JC, Bastarache L, Ritchie MD, et al. Systematic comparison of phenome-wide association study of electronic medical record data and genome-wide association study data. *Nat Biotechnol*. 2013;31(12):1102-1110. doi:10.1038/nbt.2749

2.	Wu P, Gifford A, Meng X, et al. Mapping ICD-10 and ICD-10-CM codes to phecodes: Workflow development and initial evaluation. *J Med Internet Res*. 2019;21(11):1-13. doi:10.2196/14325

3.	Zheutlin AB, Dennis J, Linnér RK, et al. Penetrance and pleiotropy of polygenic risk scores for schizophrenia in 106,160 patients across four health care systems. *American Journal of Psychiatry*. 2019;176(10):846-855. doi:10.1176/appi.ajp.2019.18091085

4.	Bigdeli TB, Voloudakis G, Barr PB, et al. Penetrance and Pleiotropy of Polygenic Risk Scores for Schizophrenia, Bipolar Disorder, and Depression among Adults in the US Veterans Affairs Health Care System. *JAMA Psychiatry*. 2022;79(11):1092-1101. doi:10.1001/jamapsychiatry.2022.2742

5.	Zhou W, Nielsen JB, Fritsche LG, et al. Efficiently controlling for case-control imbalance and sample relatedness in large-scale genetic association studies. *Nat Genet*. Published online 2018:212357. doi:10.1038/s41588-018-0184-y

6.	Yang J, Zaitlen NA, Goddard ME, Visscher PM, Price AL. Advantages and pitfalls in the application of mixed-model association methods. *Nat Genet*. 2014;46:100. doi:10.1038/ng.2876 https://www.nature.com/articles/ng.2876#supplementary-information

7.	Willer CJ, Li Y, Abecasis GR. METAL: fast and efficient meta-analysis of genomewide association scans. *Bioinformatics*. 2010;26(17):2190-2191. doi:10.1093/bioinformatics/btq340

8.	Karlsson Linnér R, Mallard TT, Barr PB, et al. Multivariate analysis of 1.5 million people identifies genetic associations with traits related to self-regulation and addiction. *Nat Neurosci*. Published online 2021:1-10. doi:10.1038/s41593-021-00908-3

9.	Metzl JM, Roberts DE. Structural competency meets structural racism: race, politics, and the structure of medical knowledge. *AMA J Ethics*. Published online 2014:674-690.

10.	Metzl JM. The Protest Psychosis &amp; the Future of Equity &amp;                    Diversity Efforts in American Psychiatry. *Daedalus*. 2023;152(4):92-110. doi:10.1162/daed_a_02033

11.	Yan C, Zhang X, Yang Y, et al. Differences in Health Professionals’ Engagement With Electronic Health Records Based on Inpatient Race and Ethnicity. *JAMA Netw Open*. 2023;6(10):e2336383-e2336383. doi:10.1001/jamanetworkopen.2023.36383

12.	de Leeuw CA, Mooij JM, Heskes T, Posthuma D. MAGMA: Generalized Gene-Set Analysis of GWAS Data. *PLoS Comput Biol*. 2015;11(4):e1004219. doi:10.1371/journal.pcbi.1004219

13.	Watanabe K, Taskesen E, van Bochoven A, Posthuma D. Functional mapping and annotation of genetic associations with FUMA. *Nat Commun*. 2017;8(1):1-11. doi:10.1038/s41467-017-01261-5

14.	Zhang MJ, Hou K, Dey KK, et al. Polygenic enrichment distinguishes disease associations of individual cells in single-cell RNA-seq data. *Nat Genet*. 2022;54(10):1572-1580. doi:10.1038/s41588-022-01167-z

15.	Friedman MJ, Huber BR, Brady CB, et al. VA’s National PTSD Brain Bank: a National Resource for Research. *Curr Psychiatry Rep*. 2017;19(10):73. doi:10.1007/s11920-017-0822-6

16.	Chatzinakos C, Pernia CD, Morrison FG, et al. Single-Nucleus Transcriptome Profiling of Dorsolateral Prefrontal Cortex: Mechanistic Roles for Neuronal Gene Expression, Including the 17q21.31 Locus, in PTSD Stress Response. *American Journal of Psychiatry*. 2023;180(10):739-754. doi:10.1176/appi.ajp.20220478

17.	Benjamini Y, Hochberg Y. Controlling the False Discovery Rate: A Practical and Powerful Approach to Multiple Testing. *Journal of the Royal Statistical Society Series B (Methodological)*. 1995;57(1):289-300. http://www.jstor.org/stable/2346101

18.	Bigdeli TB, Chatzinakos C, Bendl J, et al. Biological insights into schizophrenia from ancestrally diverse populations. *Nature*. Published online 2026. doi:10.1038/s41586-025-10000-6

19.	Colbert SMC, Group the PGCSW, Ruderfer D, Docherty AR, Mullins N. Genome-wide association studies identify 77 loci for suicidality and provide novel biological insights. *medRxiv*. Published online January 1, 2025:2025.10.22.25338076. doi:10.1101/2025.10.22.25338076

20.	Agrawal A, Brislin SJ, Bucholz KK, et al. The Collaborative Study on the Genetics of Alcoholism: Overview. *Genes Brain Behav*. 2023;22(5):e12864. doi:10.1111/gbb.12864

21.	Begleiter H. The Collaborative Study on the Genetics of Alcoholism. *Alcohol Health Res World*. 1995;19(3):228-236.

22.	Dick DM, Balcke E, McCutcheon V, et al. The collaborative study on the genetics of alcoholism: Sample and clinical data. *Genes Brain Behav*. 2023;22(5):e12860. doi:10.1111/gbb.12860

23.	Bucholz KK, Cadoret R, Cloninger CR, et al. A new, semi-structured psychiatric interview for use in genetic linkage studies: a report on the reliability of the SSAGA. *J Stud Alcohol*. 1994;55(2):149-158. doi:10.15288/jsa.1994.55.149

24.	Bucholz KK, McCutcheon V V., Agrawal A, et al. Comparison of Parent, Peer, Psychiatric, and Cannabis Use Influences Across Stages of Offspring Alcohol Involvement: Evidence from the COGA Prospective Study. *Alcohol Clin Exp Res*. 2017;41(2):359-368. doi:10.1111/acer.13293

25.	National Academies of Sciences  and Medicine E. *Using Population Descriptors in Genetics and Genomics Research: A New Framework for an Evolving Field*. The National Academies Press; 2023. doi:10.17226/26902

26.	Johnson EC, Salvatore JE, Lai D, et al. The collaborative study on the genetics of alcoholism: Genetics. *Genes Brain Behav*. 2023;22(5):e12856. doi:10.1111/gbb.12856

27.	Lai D, Kapoor M, Wetherill L, et al. Genome-wide admixture mapping of DSM-IV alcohol dependence, criterion count, and the self-rating of the effects of ethanol in African American populations. *American Journal of Medical Genetics, Part B: Neuropsychiatric Genetics*. 2021;186(3):151-161. doi:10.1002/ajmg.b.32805

28.	Harris KM, Halpern CT, Whitsel E, et al. The National Longitudinal Study of Adolescent to Adult Health: research design. *Add Health: The National Longitudinal Study of Adolescent to Adult Health*. Preprint posted online 2009. http://www.cpc.unc.edu/projects/addhealth/design

29.	Cottler LB, Robins LN, Helzer JE. The Reliability of the CIDI-SAM: a comprehensive substance abuse interview. *Br J Addict*. 1989;84(7):801-814. doi:10.1111/j.1360-0443.1989.tb03060.x
